# Cortico-cortical connectivity is influenced by levodopa in tremor-dominant Parkinson’s disease

**DOI:** 10.1101/2022.02.22.22270921

**Authors:** B.K. Rurak, J. Tan, J.P. Rodrigues, B.D. Power, P.D. Drummond, A.M. Vallence

## Abstract

**Background:** Resting tremor is the most common presenting motor symptom in Parkinson’s disease (PD). The supplementary motor area (SMA) is one of the main targets of the basal ganglia-thalamo-cortical circuit and has direct, facilitatory connections with the primary motor cortex (M1), which is important for the execution of voluntary movement. Dopamine potentially modulates SMA and M1 activity, and both regions have been implicated in resting tremor.

**Objective:** To examine SMA-M1 connectivity in individuals with PD ON and OFF dopamine medication, and whether SMA-M1 connectivity is implicated in resting tremor.

**Methods:** Dual-site transcranial magnetic stimulation was used to measure SMA-M1 facilitatory connectivity in PD participants ON and OFF levodopa. Resting tremor was measured using electromyography and accelerometry.

**Results:** Stimulating SMA had an inhibitory influence on M1 excitability OFF levodopa, and a facilitatory influence on M1 excitability ON levodopa. ON medication, correlational analysis showed an association between tremor severity and SMA-M1 connectivity, with SMA-M1 facilitation associated with smaller tremor than SMA-M1 inhibition.

**Conclusions:** The current findings contribute to our understanding of the neural networks involved in PD which are altered by levodopa medication and provide a neurophysiological basis for the development of interventions to treat resting tremor.

## Introduction

Parkinson’s disease (PD) is a heterogeneous neurodegenerative disorder characterized by motor and non-motor symptoms^1^. Tremor, involuntary and rhythmic movements of one or more body parts, is the most common presenting motor symptom and develops early in the disease^2,3^. The pathological hallmark of PD is a progressive degeneration of dopaminergic neurons in the basal ganglia, impacting the nigrostriatal pathway, resulting in altered function in subcortical and cortical motor areas^4,5^.

The supplementary motor area (SMA) is one of the main targets of the basal ganglia-thalamo-cortical circuit, receiving input from the globus pallidus indirectly via the motor thalamic nuclei^6–10^. The main efferent pathway from SMA is to the primary motor cortex (M1)^11,12^. Evidence from functional magnetic resonance imaging (fMRI) in PD shows greater blood-oxygen-level dependent (BOLD) activity in SMA and M1 ON than OFF levodopa medication during motor tasks^13,14^, suggesting that dopamine modulates both SMA and M1 activity in PD. fMRI can show brain regions simultaneously active during a task, but it is unclear whether changes in BOLD signals are due to excitation of facilitatory and/or inhibitory circuits.

Transcranial magnetic stimulation (TMS), a non-invasive brain stimulation technique, can measure interactions between regions in the cortical motor network^15–17^. Single-pulse TMS provides a measure of corticospinal excitability: a single suprathreshold TMS pulse delivered to M1 elicits a motor-evoked potential (MEP) in a target muscle. Dual-site TMS provides a measure of interactions between SMA and M1: a conditioning stimulus delivered to SMA before a test stimulus delivered to M1 with inter-stimulus intervals (ISI) of 6-7ms facilitates MEP amplitude compared to a test stimulus-alone^15,17–19^, likely due to the activation of glutamatergic excitatory interactions between SMA and M1^12,20^. Dual-site TMS reliably measures SMA-M1 connectivity in younger and older adults^19^.

Given the evidence that dopamine modulates SMA and M1 activity^13,14^, the primary aim of the current study was to examine SMA-M1 connectivity in PD ON and OFF levodopa medication. We hypothesized that SMA-M1 facilitation would be greater ON than OFF medication. Previous research has shown that (1) single-pulse TMS delivered to SMA and M1, but not the cerebellum, interrupts ongoing tremor activity and resets tremor to a new point in the tremor cycle; and (2) single-pulse TMS to M1, but not cerebellum, significantly reduces resting tremor power^21,22^. Given previous research showing disruptions in the corticospinal-motoneuronal pathway are associated with tremor, we also explored possible associations between SMA-M1 connectivity and tremor ON and OFF medication.

## Methods Participants

Eighteen people with PD recruited from a hospital outpatient clinic participated (Supplementary Table 1 shows participant characteristics). Participants were excluded if they had contraindications to TMS, potential cognitive impairments, or advanced Parkinson’s disease progression (see Supplementary Materials *S1* for screening details). All participants had a clinical diagnosis of idiopathic PD by a movement disorder neurologist (J.P.R), resting tremor involving at least one upper limb, and were treated with levodopa. Patients with head tremor or severe upper limb dyskinesia were excluded to avoid difficulties with TMS coil placement. Inclusion and exclusion criteria were assessed by a neurologist and neuropsychiatrist. The protocol was approved by Murdoch University Human Research Ethics Committee. All participants gave written informed consent.

## TMS

Participants sat with both forearms resting on chair armrests. Electromyographic (EMG) activity was recorded from the relaxed first dorsal interosseous (FDI) and extensor carpi radialis (ECR) of the most affected arm using Ag-AgCI surface electrodes in a belly-tendon montage. EMG activity was amplified (x1000; CED 1902), bandpass filtered at 1-1000Hz, notch-filtered at 50Hz, and digitized at 5kHz (CED 1401). Dual-site TMS was delivered using two figure-of-eight coils (50-mm diameter), connected to Magstim 200^2^ stimulators (Magstim Co., UK). Neuronavigation software (Brainsight TMS, Rogue Research, Canada) was used to monitor coil position.

The M1 stimulation coil was placed tangentially to the scalp with the handle positioned backwards, away from the midline by ∼45° to induce a posterior-anterior current in M1 contralateral to the affected arm (Figure 1A; *n*=4 left arm). (Detailed procedure in Supplementary Materials *S2.*) The SMA stimulation coil was placed on the midline, 4 cm anterior to the vertex^15–17^, using a lateral orientation (Figure 1B;^15–19^).

**Figure 1.**
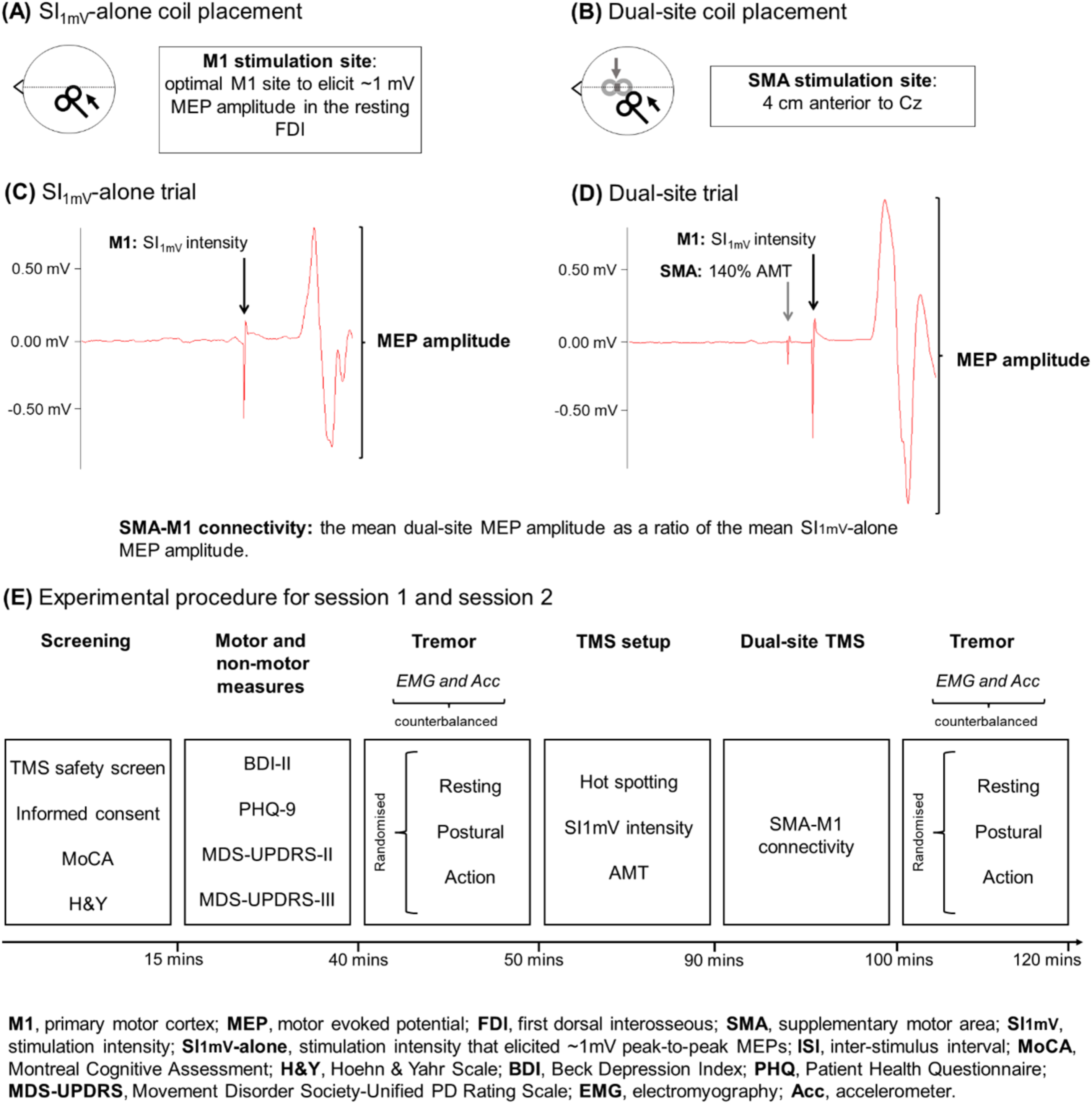
SMA-M1 connectivity was assessed by delivering SI_1mV_-alone and dual-site trials. Panel A shows the coil placement and current flow direction (indicated by the arrow) for SI_1mV_-alone trials delivered to M1, and Panel C shows an example MEP elicited by SI_1mV_-alone trials to M1 (∼1mV MEP amplitude). Panel B shows the coil placement for dual-site trials: the grey coil represents the SMA stimulation site, and the black coil represents the M1 stimulation site, with arrows indicating current flow direction. Panel D shows an example MEP amplitude elicited by dual-site TMS which involves delivering a conditioning stimulus to SMA (140% AMT) 7 ms before a test stimulus to M1. All measures were obtained from the tremor-affected limb. EMG and tri-axial accelerometer measures of tremor were counterbalanced across sessions and participants. SMA-M1 connectivity was quantified as the mean dual-site MEP amplitude as a ratio of the mean SI_1mV_-alone MEP amplitude. Panel E shows the experimental procedure for ON and OFF medication: experimental sessions were separated by a minimum of 7 days.

## Stimulation intensities

Consistent with all previous reports using dual-site TMS to measure SMA-M1 connectivity^15–19^, M1 TMS intensity was set as the intensity (as a percentage of maximum stimulator output; %MSO) that elicited peak-to-peak MEP amplitudes of ∼1mV in the resting FDI (SI_1mV_), and SMA stimulation intensity was set as 140% of active motor threshold (AMT). AMT was defined as the minimum intensity (%MSO) that elicited MEPs in FDI ≥0.2mV from at least 5/10 consecutive trials during an isometric contraction of 10% maximum voluntary contraction^23^. TMS intensities were determined in both sessions. MEP amplitude elicited from single-pulse trials using SI_1mV_ delivered to M1 was defined as ‘SI_1mV_-alone’ MEP amplitude (Figure 1C). There were no differences in TMS parameters between sessions (see Supplementary Materials *S2.1* and *S2.2*).

## Experimental Protocol

Figure 1E shows the experimental procedure. Each participant completed two 2-hour experimental sessions: (1) ON, defined as starting the experimental session ∼1hr after taking levodopa (range: 60-75 minutes); (2) OFF, defined as starting the experimental session ≥12hrs after overnight withdrawal from levodopa (range: 12-15.5 hours). The sessions were counterbalanced across participants, separated by ≥7 days (first session on average 7.67±1.45 days before the second session), and completed in the morning (range: 6:00-10:30 a.m.) to coincide with individual dosing times and allow ≥12hrs medication withdrawal.

## SMA-M1 connectivity

SMA-M1 connectivity was assessed using SI_1mV_-alone (Figure 1C) and dual-site trials (Figure 1D). For dual-site trials, a conditioning pulse delivered to SMA (140% AMT) preceded a test pulse delivered to M1 (SI_1mV_) by an ISI of 7 ms^19^. SI_1mV_-alone and dual-site trials were pseudo-randomized with an inter-trial interval of 5 s (±10%). TMS application was controlled using a custom-developed Signal script: the script triggered TMS pulses if EMG activity was below an individually determined EMG activity threshold for a duration of 50 ms (Figure 2A). The individual EMG activity thresholds were determined from EMG activity recorded for the measurement of resting tremor before TMS was administered (see “Measures of PD tremor” section for details of tremor measurements). If EMG activity was not below the EMG activity threshold for 50 ms, TMS triggered after ∼5 s (±2%), indicating some EMG activity at the time of TMS delivery (Figure 2B): these trials were excluded from the analysis. TMS was triggered after 5 s to limit the length of the experimental session. See Supplementary Materials section *S3* for details of the script and individual EMG thresholds.

**Figure 2.**
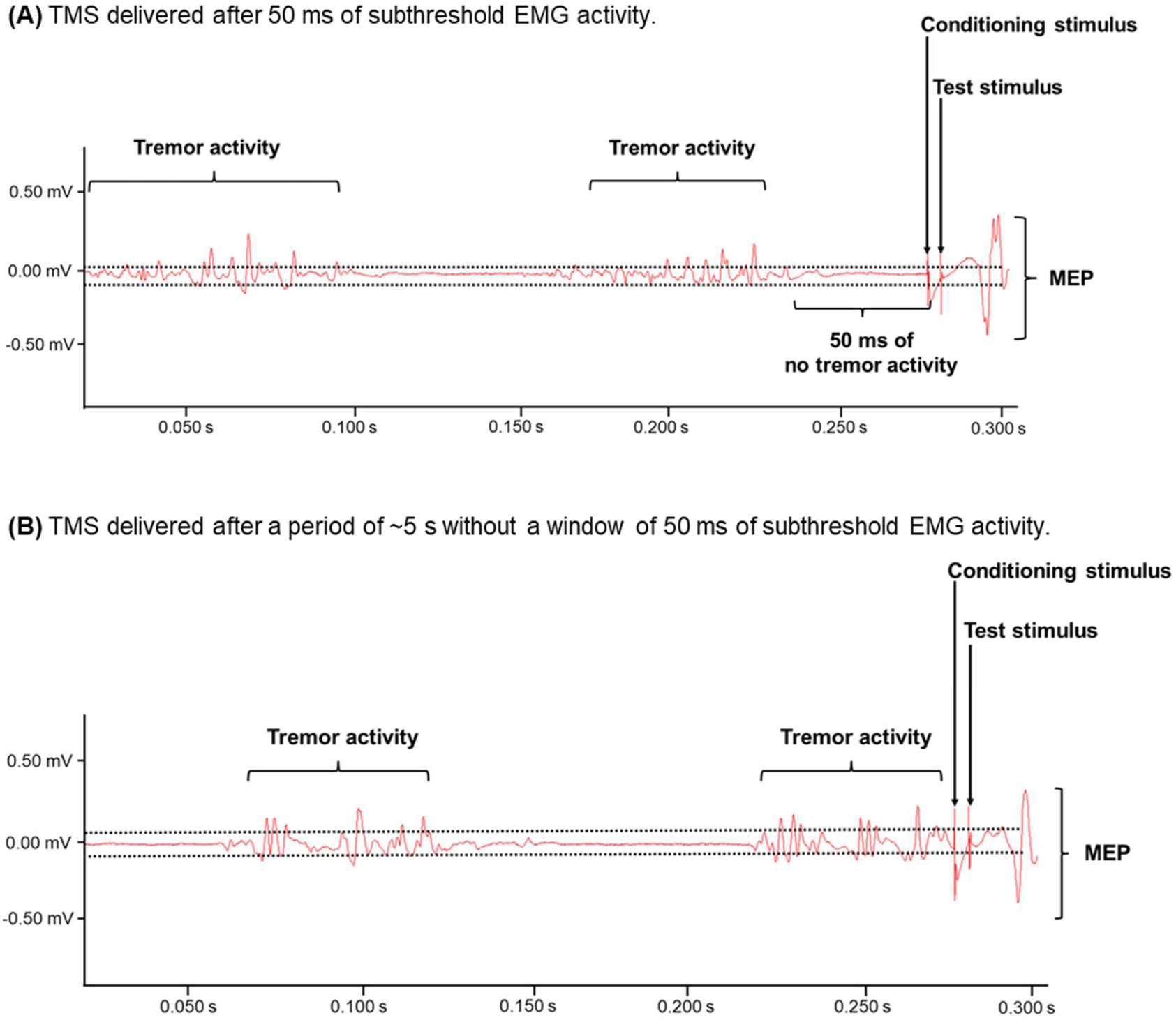
Example EMG traces from the data acquisition phase (50-Hz notch filter, 1 Hz high pass filter, 1000 Hz low pass filter) for TMS trials from a participant OFF medication. SI_1mV_-alone and dual-site trials were delivered after 50 ms of EMG activity below an EMG activity threshold determined for each participant (A) or if a period of ∼5 s passed without EMG activity below the determined EMG activity threshold (B; these trials were removed from analysis). (B) shows the last 0.300 s of the 5-s period. The two horizontal dotted lines reflect the predetermined threshold; EMG activity within these lines indicates EMG activity below the tremor threshold. Thresholds (i.e., peak amplitudes; mV) were established for each participant before TMS.

Two experimental blocks each consisting of 30 trials were delivered: 15 SI_1mV_-alone trials and 15 dual-site trials. Blocks lasted ∼4 minutes with a 1-2-minute break between blocks. Participants were instructed to remain quiet, not suppress tremor activity, keep their eyes open, and stay alert during TMS^24,25^. Although stimulation parameters were optimized for the FDI, MEPs were simultaneously recorded from FDI and ECR and SMA-M1 connectivity was quantified separately from FDI and ECR MEPs.

## Measures of PD tremor

Resting, postural and action tremor were measured using the Movement Disorder Society Unified Parkinson’s Disease Rating Scale (MDS-UPDRS), a tri-axial accelerometer, and EMG from the most-affected upper limb. Resting tremor measures are reported here and postural and action tremor measures are reported in Supplementary Materials section *S5*. For resting tremor, participants were instructed to relax their forearm on the chair armrest without their hand touching anything. MDS-UPDRS was assessed at the beginning of each session, ∼35 minutes before TMS setup began. EMG and accelerometer measures of tremor were assessed ∼5 minutes before and ∼5 minutes after TMS blocks to identify whether tremor changed throughout the experimental session.

## MDS-UPDRS

MDS-UPDRS part III items 17 and 18 were used to assess the severity of resting tremor using a 5-point scale (0: normal, 1: slight, 2: mild, 3: moderate, 4: severe)^1^. The MDS-UPDRS was rated by a certified MDS-UPDRS assessor.

## EMG tremor

Surface EMG was used to measure the muscular activity involved in tremor and was recorded from the FDI, ECR and flexor carpi radialis (FCR) using Ag-AgCI surface electrodes in a belly-tendon montage. The EMG signal was amplified (x1000; CED 1902), bandpass filtered (1-500Hz), notch-filtered (50Hz), and digitized at 5kHz (CED 1401). After data acquisition, the EMG signal was down-sampled (1000Hz), rectified, and bandpass filtered (second-order Butterworth filter; 1-30Hz) to analyse tremor activity.

## Acceleration tremor

A tri-axial accelerometer (Arduino GY-61 ADXL335; length: 21mm; width: 15mm; height 11mm) was used to measure tremor changes in acceleration in x, y, and z planes and was secured to the distal phalanx of the index finger with tape. Acceleration in three dimensions was amplified (x1000; CED 1902), bandpass filtered (2-30Hz) and digitized at a sampling rate of 5kHz (CED 1401). After data acquisition, the accelerometry signal was rectified and bandpass filtered (3-10Hz) to analyse tremor activity. Technical issues prevented acceleration measurement for two participants.

## Data analysis

All analyses were performed using R (version 4.3.2). Shapiro-Wilk’s tests for normality showed MEP and tremor data were not normally distributed. Generalized linear mixed effect models (GLMMs) were therefore used to analyse MEP and tremor data. As both MEP and tremor data were non-negative and positively-skewed, a gamma distribution with a log link function was applied for each GLMM^26^. Wald Chi-Squared tests were conducted for null hypothesis significance testing of main and interaction effects. Significant effects were investigated with Bonferroni-corrected contrasts. Effect sizes were reported as Cohen’s *d*, with *d*≤0.2 for a small effect, 0.2<*d* ≤ 0.5 for a medium effect, and *d*≥0.8 for a large effect^27^. Statistical significance was set at *α*=0.05.

## Resting tremor

For the acceleration data, a principal components analysis was performed to identify the most dominant tremor acceleration axis on which to perform the power spectral density analysis. For both the acceleration and EMG data, Welch’s power spectral density analysis was performed to identify the power of the tremor peak frequency for each individual, which was used for subsequent analyses.

Paired-sample *t*-tests were performed to examine differences in MDS-UPDRS resting tremor severity scores ON and OFF medication. GLMMs were used to analyse tremor power (separate models for EMG and acceleration tremor measures), with fixed factors of Muscle (FDI, ECR, FCR), Medication state (ON, OFF), and Time (before-TMS, after-TMS), with by-subject intercept as a random effect.

## Neurophysiological measures

Trials with EMG activity (root mean square; RMS) from the 50 ms immediately preceding TMS (pre-TMS RMS) >0.2 mV were excluded from all analyses (total n=2 trials). Equivalence testing using the two-sided statistical tests (TOST) approach^28^ was performed to determine whether pre-TMS RMS in ON and OFF medication states for SI_1mV_-alone and dual-site trials differed (separate tests for FDI and ECR). Wilcoxon signed-rank tests were used for the TOST, and the equivalence bound was based on a smallest effect size of interest of Cohen’s *d*_z_ = 0.30^29^. Single-trial-level MEP amplitudes were analysed with a GLMM with fixed factors of Trial-type (SI_1mV_-alone, dual-site), Muscle (FDI, ECR), and Medication state (ON, OFF), with by-subject intercept as a random effect. To account for the effect of EMG background activity on MEP amplitudes, pre-TMS RMS was included as a covariate in the GLMM.

## SMA-M1 Connectivity Between PD and Controls

Analyses were performed to examine differences in MEP amplitudes between the PD participants OFF medication, age– and sex-matched controls (n=14), and younger controls (n=30). The control data were from a previous study that examined SMA-M1 connectivity in younger and older adults: SMA-M1 connectivity was measured using the same TMS parameters reported in the current study^19^. As the control data were only obtained from FDI, only FDI data for the PD participants were used in this analysis. Although identical stimulation parameters were used to measure SMA-M1 connectivity in PD and control samples, other aspects of the experimental procedures differed between the two studies. For example, the UPDRS and tremor measures were conducted in PD but not controls, and dexterity and additional connectivity measures were conducted in controls but not PD.

MEP amplitudes were analysed with a GLMM with fixed factors of Group (PD-OFF, age– and sex-matched controls, younger controls), and Trial-type (SI_1mV_-alone, dual-site), with by-subject intercept as a random effect, and pre-TMS RMS as a covariate.

## Relationship between resting tremor and SMA-M1 connectivity

Exploratory correlation analyses were performed using Spearman’s rank correlation coefficient (ρ) to examine the potential relationship between SMA-M1 connectivity (FDI, ECR) and tremor measures (EMG, acceleration). The *p*-values were adjusted with the false discovery rate (FDR) to control for Type I error^30^. SMA-M1 connectivity was quantified by expressing the mean dual-site MEP amplitude as a ratio of the mean SI_1mV_-alone MEP amplitude. Ratios >1.0 indicate a facilitatory effect of SMA stimulation on M1 excitability; ratios <1.0 indicate an inhibitory effect of SMA stimulation on M1 excitability. For EMG tremor measures, associations were performed for FDI SMA-M1 connectivity and EMG recorded from FDI (before– and after-TMS), and ECR SMA-M1 connectivity and EMG recorded from ECR (before– and after-TMS).

## Results Resting tremor

A paired-sample *t*-test showed a significant difference in MDS-UPDRS resting tremor severity scores ON and OFF medication (*t*_17_=-3.25, *P*=0.005, *d*=0.77). The GLMM analysis on EMG resting tremor power found a significant main effect of Medication (χ^2^(1, N=18) =28.19, *P*<0.0001), with significantly higher tremor power OFF medication. No other significant main effects or interaction effects were found (χ^2^s<3.40, *Ps*>0.065). The GLMM analysis on resting tremor power measured with acceleration found no significant main effects or interaction effects (χ^2^s<3.14, *Ps*>0.076).

## SMA-M1 connectivity

Equivalence tests found that pre-TMS RMS was statistically equivalent when comparing ON and OFF medication state for FDI during SI_1mV_-alone (*V*=79, *P*=0.603) and dual-site trials (*V*=103, *P*=0.783), and for ECR during SI_1mV_-alone (*V*=87, *P*=0.535) and dual-site trials (*V*=108, *P*=0.842).

The GLMM analysis on MEP amplitudes found a significant Trial-type X Medication X Muscle X pre-TMS RMS interaction (χ^2^(1, N=18)=4.75, *P*=0.029). Figure 3 shows FDI and ECR MEP amplitudes for SI_1mV_-alone and dual-site trials ON and OFF medication. Post-hoc analyses showed that OFF medication, MEP amplitudes from dual-site trials were significantly smaller than SI_1mV_-alone trials for both FDI (*z*=-2.06, *P*=0.039, *d*=-0.15) and ECR muscles (z=-2.35, *P*=0.019, *d*=-0.17), indicating an inhibitory effect of SMA stimulation on M1 excitability. By contrast, ON medication, MEP amplitudes from dual-site trials were significantly larger than SI_1mV_-alone trials for FDI (z=2.16, *P*=0.031, *d*=0.16) and ECR muscles (z=2.45, *P*=0.014, *d*=0.18), which indicates a facilitatory effect of SMA stimulation on M1 excitability. See Supplementary Materials section *S6* for the effects of pre-TMS RMS as a covariate.

**Figure 3.**
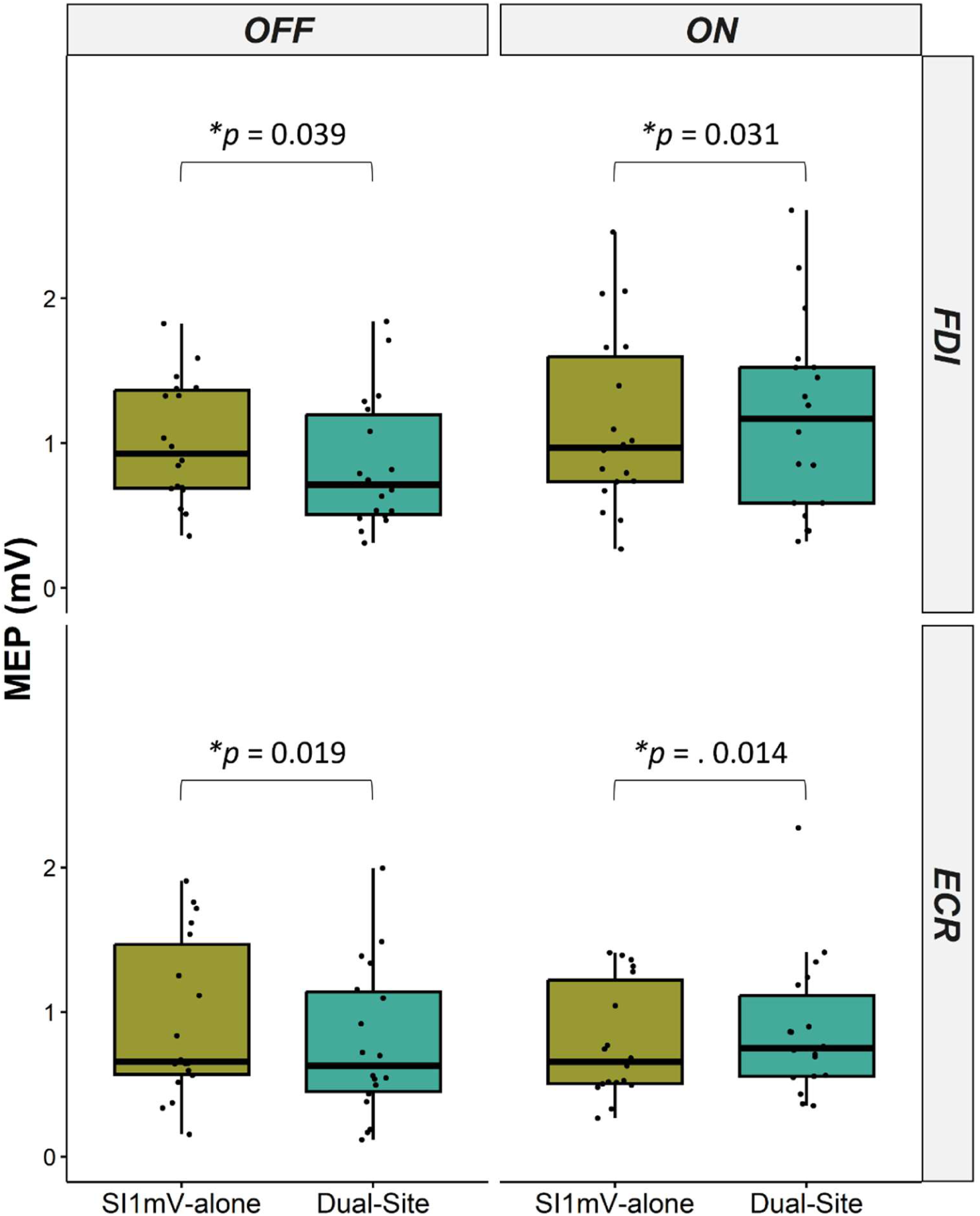
MEP amplitudes from FDI and ECR muscles, for SI_1mV_-alone and dual-site trials, ON and OFF medication. OFF medication, MEP amplitudes from FDI and ECR from dual-site trials were significantly smaller than SI_1mV_-alone trials, which indicates an inhibitory effect of SMA stimulation on M1 excitability. By contrast, ON medication, MEP amplitudes from FDI and ECR from dual-site trials were significantly larger than SI_1mV_-alone trials, which indicates a facilitatory effect of SMA stimulation on M1 excitability. MEP amplitudes from FDI and ECR from dual-site trials were also significantly smaller OFF medication compared to ON medication. *\*P < 0.05*.

The GLMM analysis also found a significant main effect of Muscle (χ^2^(1, N=18)=6.82, *P*=0.009), with MEP amplitudes from FDI significantly larger than MEPs from ECR. This is expected as the stimulation intensities and site were optimized for FDI, and TMS thresholds roughly follow a proximo-distal gradient, with lower thresholds in more distal muscles.

## SMA-M1 Connectivity Between PD and Controls

Figure 4 shows the MEP amplitudes from SI_1mV_-alone and dual-site trials for PD OFF medication, age– and sex-matched controls, and younger controls. There was a significant Group X Trial-type interaction (χ^2^(1, N=62)=10.02, *P*=0.0067. Follow-up analyses replicated the inhibitory effect of SMA stimulation on M1 excitability for PD OFF medication, with MEP amplitudes from dual-site trials significantly smaller than SI_1mV_-alone trials (*z*=-3.25, *P*=0.001, *d*=-0.24). By contrast, there was no significant difference in MEP amplitudes from dual-site trials and SI_1mV_-alone trials for age– and sex-matched controls (z=-1.36, *P*=0.173, *d*=-0.17). For younger controls, MEP amplitudes were larger for dual-site trials compared to SI_1mV_-alone trials, but this facilitatory influence of SMA stimulation on M1 excitability was not statistically significant (z=1.66, *P*=0.098, *d*=0.19). There were no other significant main and interaction effects (χ^2^s<1.37, *Ps*>0.242).

**Figure 4.**
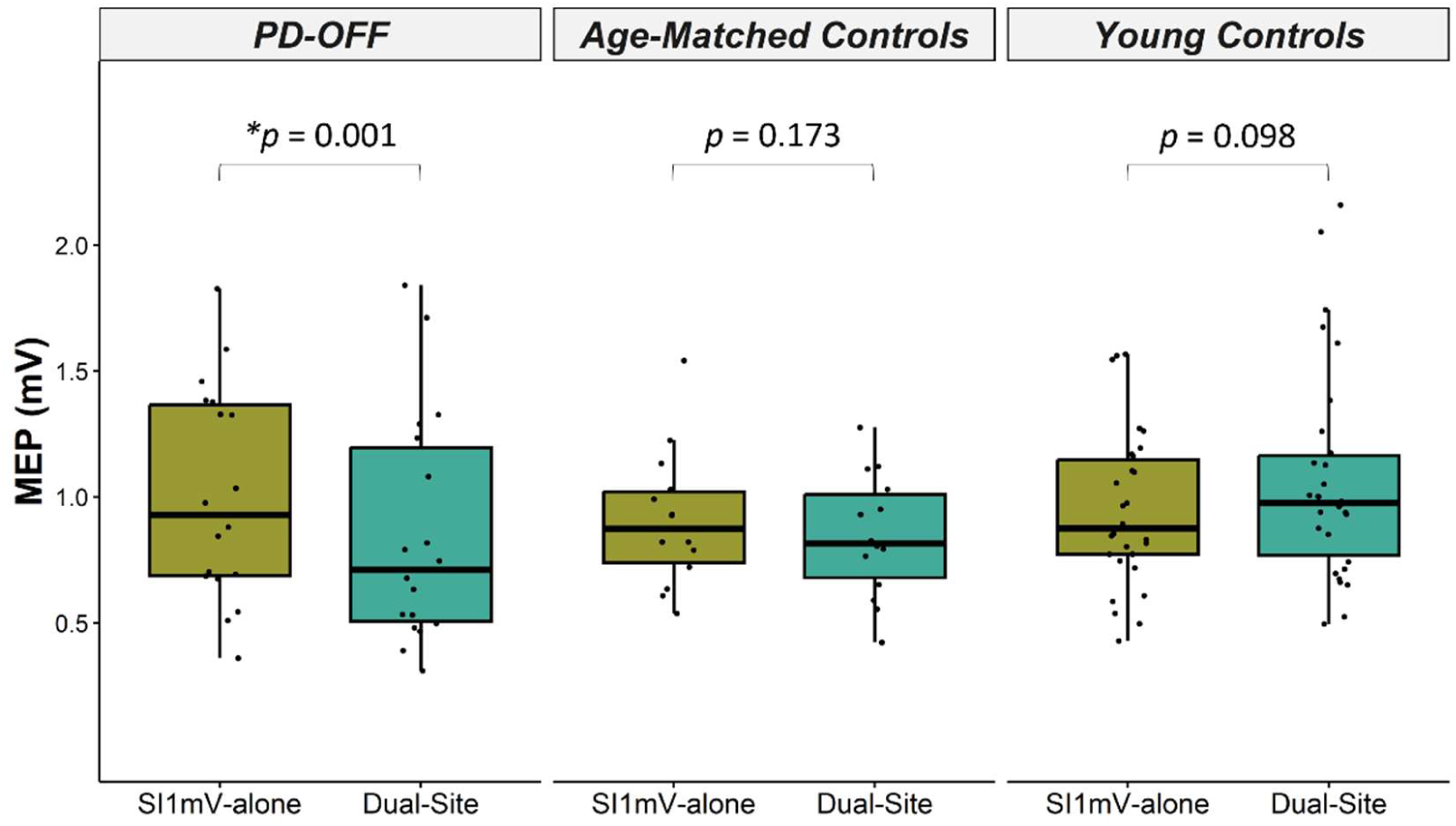
MEP amplitudes (from FDI) from SI_1mV_-alone and dual-site trials for PD OFF medication, age– and sex-matched controls, and younger controls. SMA-M1 connectivity was inhibitory in PD OFF medication, with MEP amplitudes being significantly smaller for dual-site trials compared to SI_1mV_-alone trials. *\*P < 0.05*.

## SMA-M1 connectivity and tremor

Figures 5 and 6 show associations between FDI SMA-M1 connectivity ratios and resting tremor power measured using accelerometry and EMG, respectively. ON medication, the more facilitatory FDI SMA-M1 connectivity, the less severe the tremor in FDI recorded using EMG (Figure 5C and 5D) and acceleration (Figure 6C and 6D) before and after TMS. Positive correlations between SMA-M1 connectivity and tremor were also observed in ECR, however, these associations were not significant after FDR adjustment (scatterplots showing ECR SMA-M1 connectivity and resting tremor are presented in Supplementary Materials section *S6*). No other significant associations were found between SMA-M1 connectivity and resting tremor. Scatterplots showing FDI and ECR SMA-M1 connectivity and postural and action tremor are presented in Supplementary Materials section *S7*.

**Figure 5.**
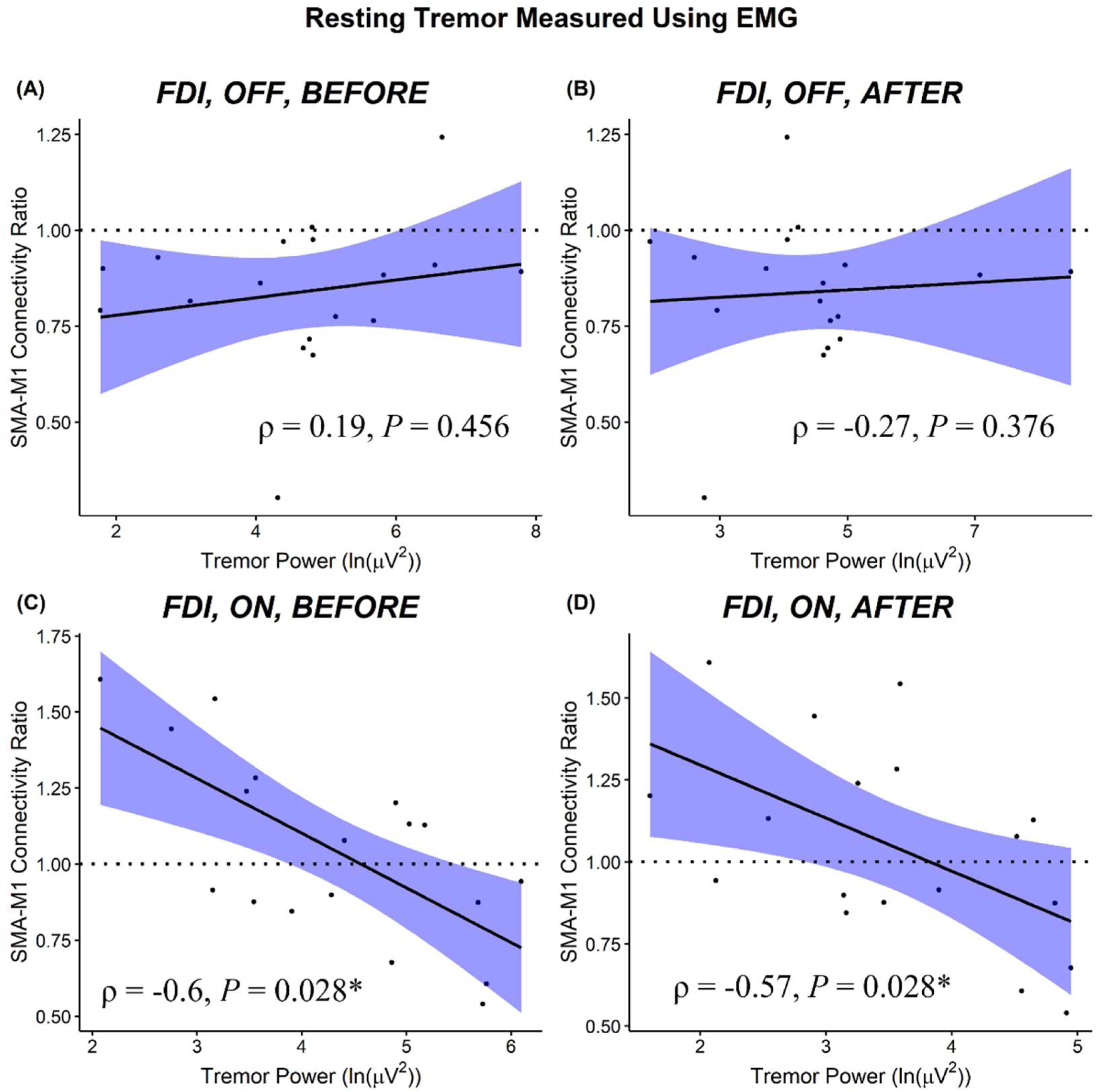
Scatterplots show the relationship between the magnitude of FDI SMA-M1 connectivity ratios and FDI resting tremor power (ln(µV^2^)) measured using EMG. ON medication, SMA-M1 connectivity that was more facilitatory (>1) was significantly associated with lower tremor power. 95% confidence interval bands are shown. \**P* < 0.05.

**Figure 6.**
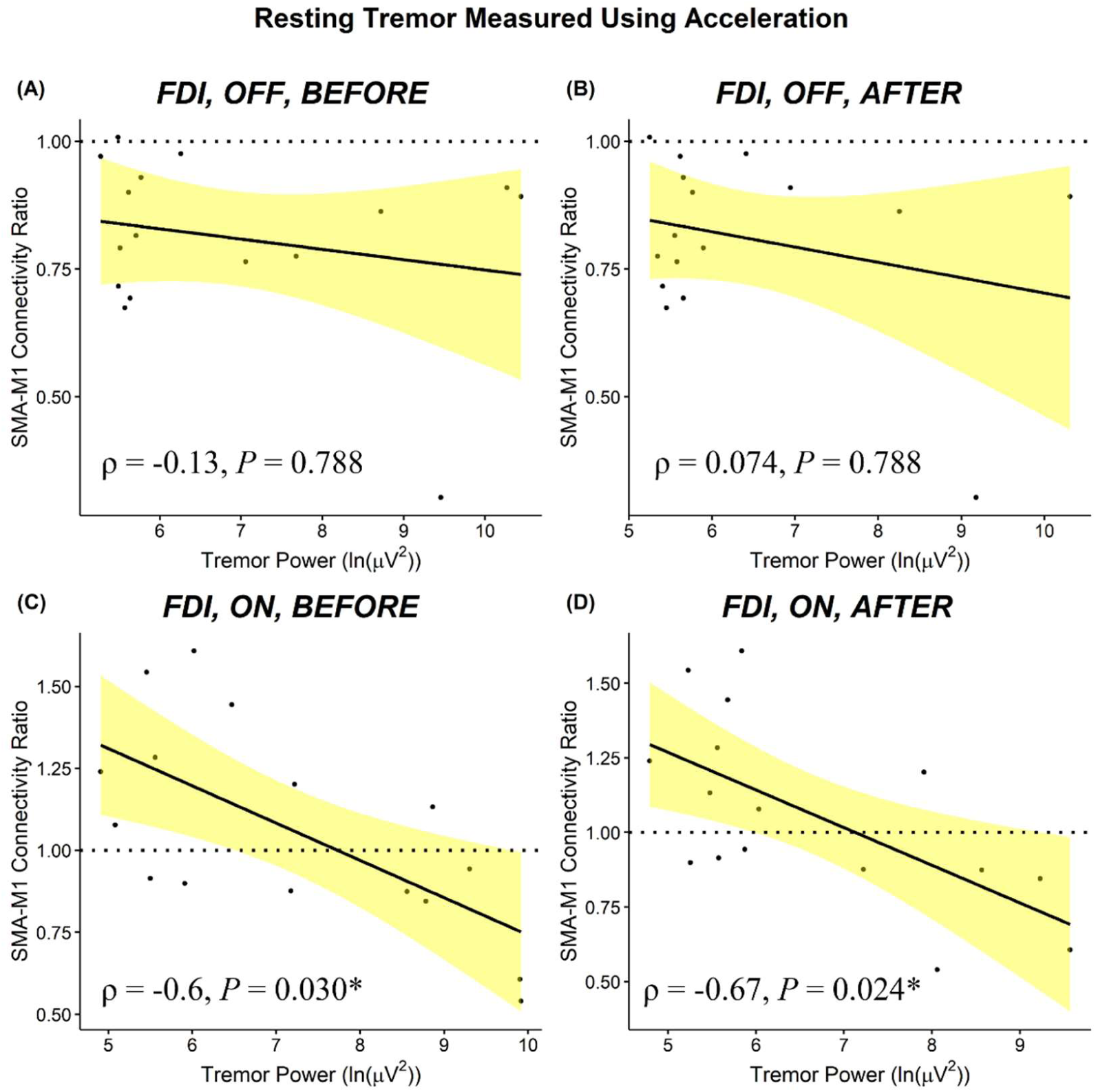
Scatterplots show the relationship between the magnitude of FDI SMA-M1 connectivity ratios and FDI resting tremor power ((ln(µg^2^))) measured using acceleration. ON medication, SMA-M1 connectivity that was more facilitatory (>1) was significantly associated with lower tremor power. 95% confidence interval bands are shown. \**P* < 0.05.

## Discussion

We examined SMA-M1 connectivity using dual-site TMS in people with PD ON and OFF levodopa medication. There was an inhibitory influence of SMA stimulation on M1 excitability OFF medication in 88% of participants. Conversely, SMA stimulation facilitated M1 excitability ON medication in 50% of participants. Furthermore, correlational analysis showed an association between tremor severity and SMA-M1 connectivity, with SMA-M1 facilitation associated with smaller tremor than SMA-M1 inhibition.

## OFF medication: SMA-M1 inhibition

Trial-level analysis showed that MEP amplitude was significantly smaller for dual-site stimulation targeting SMA and M1 compared to M1 stimulation alone, suggesting a significant inhibitory influence of SMA stimulation on M1 excitability. The inhibitory influence of SMA stimulation on M1 excitability in PD OFF medication likely reflects disease-related alterations in the basal ganglia-thalamo-cortical circuit because of a loss in dopaminergic cells. According to the classical “rate model”, loss of dopamine in PD is thought to elicit an abnormal increase in inhibitory drive from the globus pallidus internal to the thalamus^5^. In turn, the faciliatory projection from the motor thalamus to the cerebral cortex is thought to be suppressed, resulting in more inhibitory than faciliatory activity in the cerebral cortex^5^. Thus, dopamine-related changes in the basal ganglia-thalamo-cortical circuit might preferentially activate inhibitory connections between SMA-M1 and, in part, underpin the SMA-M1 inhibition found in the current study. However, this is speculative, and the exact role of this circuit remains elusive.

SMA-M1 inhibition found in PD OFF medication might also be due to alterations in the cerebello-thalamo-cortical circuit, which is implicated in tremor-dominant PD^31–33^. The cerebellum has anatomical connections to both SMA and M1 via the motor thalamic nuclei^7,34^. A diffusion tensor MRI study showed reduced neural transmission along the cerebello-thalamo-cortical white matter tract in tremor-dominant PD OFF medication compared to healthy controls^35^. fMRI research has shown increased cerebellar BOLD activity in PD OFF medication compared to healthy controls, suggesting hyperactivity within the cerebellum of PD^36,37^. These findings fit with dual-site TMS research showing atypical cerebellar facilitation of M1 excitability in PD OFF medication compared to healthy controls^38,39^. It is worth noting, however, that moderate and poor test-re-test reliability of dual-site TMS measures of cerebellar brain inhibition has been reported in younger and older adults, respectively^40,41^, and reliability has not been established in PD. Although speculative, alterations in the cerebello-thalamo-cortical circuit might influence the thalamic inhibitory drive and, subsequently, influence SMA excitability, underpinning the observed SMA-M1 inhibition OFF medication.

Altered SMA activity from motor thalamic nuclei (targeted by the basal ganglia and cerebellum) might influence facilitatory and inhibitory intracortical circuits within M1. Results from a triple-pulse TMS study in healthy younger adults showed that a conditioning stimulus to SMA had no influence on the excitability of short-interval intracortical inhibitory (SICI) circuits in M1 but increased short-interval intracortical facilitation (SICF) in M1 when compared to TMS given to M1-alone. These findings suggest that SICF, which reflects the nett result of a complex descending corticospinal volley comprising direct and indirect waves^42^, might contribute to the facilitatory effect of SMA stimulation on M1 excitability. No research has investigated SICF and SICI in tremor-dominant PD OFF medication, and it remains unclear whether the excitability within these circuits is altered compared to healthy controls. However, TMS research examining intracortical excitability in different PD subtypes (e.g., akinetic-rigid symptoms, levodopa-induced dyskinesia, drug-naïve individuals) has shown increased SICF^43,44^ and reduced SICI^45,46^ in PD OFF medication compared to controls. It is difficult to draw conclusions on these previous studies because tremor-dominant PD likely has a different pathophysiology to other PD subtypes^47^; however, based on the abovementioned studies, it is unlikely that SMA-M1 inhibition found in the current study was mediated by intracortical processes in M1.

## Differential effect of SMA stimulation on M1 excitability ON compared to OFF medication

SMA stimulation had an inhibitory influence on M1 excitability OFF medication and a facilitatory influence on M1 excitability ON medication. In healthy young adults, several reports show that SMA stimulation has a facilitatory influence on M1 excitability and, therefore, the current results suggest that, at the group level, levodopa normalizes SMA-M1 connectivity. Evidence from photon emission tomography research showed increased SMA cerebral blood flow activity ON compared to OFF medication in PD during simple motor tasks^48,49^. Similarly, fMRI research has shown increased BOLD activity in both the SMA and M1 ON compared to OFF medication while performing simple motor tasks^13,14^. This is thought to be due to levodopa normalising cortical activity: levodopa increases dopamine levels in the basal ganglia, which reduces nett inhibition of the motor thalamic nuclei and increases excitation of the cerebral cortex^5,50^. Our findings show a similar trend to neuroimaging research; however, it is difficult to make direct comparisons as these studies investigated non-tremor-dominant PD (i.e., akinetic PD) and investigated the effects of medication on cortical activity during motor tasks rather than at rest.

Given the main analysis examining the effect of SMA stimulation on M1 excitability in PD ON and OFF medication showed dual-site MEP amplitudes were significantly larger than SI_1mV_-alone MEP amplitudes ON medication but significantly smaller than SI_1mV_-alone MEP amplitudes OFF medication, it is tempting to speculate that dopaminergic function plays a role in SMA-M1 connectivity. Dopamine-related changes in M1 might heighten inhibitory processes and, in part, mediate SMA-M1 inhibition. Previous MRI research has primarily investigated M1 activity in non-tremor-dominant PD (e.g., akinetic, bradykinesia): M1 BOLD activity was increased during simple motor control tasks in PD OFF medication compared to healthy controls (e.g.,^14,36,51^). Furthermore, MRI research in PD has shown that M1 BOLD activity was increased ON compared to OFF medication at rest^52^ and during movement^14^. Less is known about M1 activity in tremor-dominant PD, but one fMRI study has shown increased M1 BOLD activity in tremor-dominant compared to non-tremor-dominant PD^53^; however, 12 of the 15 tremor-dominant participants did not use any PD-related medication, including levodopa, which makes it difficult to draw conclusions about M1 activity ON and OFF medication in tremor-dominant PD. Together, these findings provide some evidence to suggest that M1 activity is altered OFF levodopa medication in PD, but it is unclear whether altered M1 activity results in increased inhibitory processes that could influence SMA-M1 inhibition.

We also conducted comparisons between SMA-M1 connectivity in tremor-dominant PD OFF medication and a control sample of age– and sex-matched participants and younger participants for whom SMA-M1 connectivity data were collected in a previous study^18^. We found SMA-M1 inhibition in PD OFF medication, but not in age– and sex-matched controls. This is in line with the proposed role of dopaminergic function in SMA-M1 connectivity: both older adults and people with PD show a decline in dopaminergic neurons^4,54,55^, however, the magnitude and rate of dopamine loss is greater in PD than in older adults^3^. While the control analysis did not find significant SMA-M1 facilitation in younger controls, several previous studies have found SMA-M1 facilitation measured using dual-site TMS in younger adults^3–6^. Based on the results of this analysis, we speculate that ON medication, dopaminergic function in PD is similar to younger adults, which might explain the facilitatory interaction between SMA and M1 observed ON medication in PD participants. However, given that the control data were collected from a previous study, and with a relatively smaller sample of age– and sex-matched controls, it is important for future research to examine SMA-M1 connectivity using dual-site TMS in a large sample of older and younger adults and PD to examine the potential differences in SMA-M1 connectivity between these groups. If SMA-M1 connectivity differs significantly between the groups, this will provide the foundation to examine the role of dopamine neuron loss in SMA-M1 connectivity. It might be hypothesized that SMA-M1 inhibition is mediated by a decline in dopamine neurons, and dual-site TMS might be an important tool to monitor changes in SMA-M1 connectivity and, indirectly, dopamine neurons.

## Relationship between SMA-M1 connectivity and resting tremor

ON medication, a facilitatory influence of SMA stimulation on M1 excitability was associated with reduced tremor power, whereas an inhibitory influence of SMA stimulation on M1 excitability was associated with increased tremor power (measured using both EMG and accelerometry from FDI). Therefore, it is possible that the magnitude of SMA-M1 facilitation might play a role in modulating tremor activity. Future research should examine associations between tremor amplitude and SMA-M1 connectivity with a larger sample of individuals and a broader range of tremor severity, as well as examine whether altering SMA-M1 connectivity (potentially using cortical paired associative stimulation or transcranial electrical stimulation) affects tremor severity. The future investigation of SMA-M1 connectivity in akinetic-rigid (non-tremor dominant) PD individuals will also offer important insight into the relationship between SMA-M1 connectivity and tremor.

When correcting for multiple comparisons, there were no significant associations between SMA-M1 connectivity measured from ECR and tremor amplitude measured either using EMG or accelerometry. This might be due to the stimulation parameters being optimized for FDI, and not ECR; it would be interesting to determine whether optimising stimulation parameters for ECR reveals a relationship with both measures of tremor (EMG, acceleration).

No significant associations were found between SMA-M1 connectivity and tremor severity OFF medication. A previous fMRI study showed increased functional connectivity in SMA and M1 activity in tremor-dominant PD OFF medication compared to healthy controls, and the increase in M1 but not SMA activity was associated with increased clinical resting tremor scores^56^. These findings suggest that alterations in SMA activity OFF medication does not influence resting tremor severity. Our findings, in part, extend this previous research by showing that the excitatory influence of SMA stimulation on M1 excitability OFF medication was not associated with PD resting tremor severity. It would be interesting to examine SMA-M1 connectivity (ON and OFF medication) in tremor-dominant PD and non-tremor dominant PD to clarify whether alterations in SMA-M1 connectivity is specific to PD tremor.

## Limitations

SMA stimulation was delivered 4 cm anterior to Cz, consistent with previous research^15–19^. However, it is possible that this placement could be sub-optimal for SMA stimulation and target pre-SMA in some individuals^57^. It is important for future research to use individual anatomical scans and neuronavigation to determine the optimal SMA stimulation site. Another limitation to the study is that the OFF state was defined as a withdrawal from levodopa for a minimum of 12hrs (range: 12-15.5hrs), consistent with previous research^58,59^. Some individuals were taking long-acting dopaminergic agonist medication (*n*=8), which requires a withdrawal period of ≥72hrs^60^. Future studies should compare tremor-dominant PD participants on long-acting dopamine agonist medications with tremor-dominant PD participant not on long-acting dopamine agonist medications.

## Conclusions

This is the first study to characterize SMA-M1 connectivity measured using dual-site TMS in people with PD ON and OFF levodopa medication. Findings from this study suggest that levodopa medication alters SMA-M1 connectivity ON compared to OFF medication: the inhibitory influence of SMA stimulation on M1 excitability found OFF medication was reduced ON medication. ON medication, correlational analysis showed an association between tremor severity and SMA-M1 connectivity, with SMA-M1 facilitation associated with smaller tremor than SMA-M1 inhibition.

## Data availability

The data supporting the findings of this study are available from the corresponding author upon reasonable request.

## Acknowledgments

This study was performed at Murdoch University, Western Australia, Australia. B.K.R., J.P.R., B.D.P., P.D.D., and A.M.V conceived and designed the experiment; B.K.R. performed the experiments; B.K.R., J.T., P.D.D. and A.M.V. analysed the data; B.K.R. drafted the manuscript; P.D.D., J.T., and A.M.V. critically revised the manuscript; J.P.R., B.D.P., P.D.D., and A.M.V. provided supervision. All authors have approved the final version of the manuscript and agree to be accountable for all aspects of the work in ensuring that questions related to the accuracy or integrity of any part of the work are appropriately investigated and resolved. All persons designated as authors qualify for authorship, and all those who qualify for authorship are listed.

B.K.R. was supported by an Australian Government Research Training Program scholarship and the Graduate Women (WA) Inc. Education Trust – Barbara Mary Hale Bursary. A.M.V. was supported by an Australian Research Council Discovery Early Career Researcher Award (DE190100694). Publication of this work was supported by the Bryant Stokes Neurological Research Foundation.

