## Supplementary Materials for "Cortico-cortical connectivity is influenced by levodopa in tremor-dominant Parkinson’s disease"

### S1. Screening

Supplementary Table 1 shows participant characteristics; ages and duration of disease are presented in range to limit identification of participants. All participants completed a 17-item safety questionnaire, which was used to screen and exclude individuals with contraindications to TMS ^1-4^. The Montreal Cognitive Assessment was used to screen and exclude individuals with potential cognitive impairments; no participant scored less than the cut-off score of 25 (median 28, score range: 26-30) ^5^. The Hoehn and Yahr scale was used to screen individuals with advanced Parkinson’s disease progression, as it would be difficult for these individuals to temporarily withdraw from their dopaminergic medication; no participant received a score of 4 or greater, which would have indicated advanced Parkinson’s disease progression.

*Supplementary Table 1.*

*Demographic and clinical characteristics of participants.*

| **Participant** | **Age**  **(years)*** | **Sex** | **Disease duration in years*** | **H&Y score** | **Levodopa medication** | **Levodopa dose per day** | **Other medication** | **Levodopa equivalent daily dose^1^** | **MDS-UPDRS part III^2^** | |
| --- | --- | --- | --- | --- | --- | --- | --- | --- | --- | --- |
|  |  |  |  |  |  |  |  |  | *ON* | *OFF* |
| 1 | 70-75 | M | 10-15 | 2 | 250 mg | 4 |  | 1000 | 3 | 2 |
| 2 | 60-65 | M | 5-10 | 1 | 200 mg | 3 |  | 600 | 1 | 2 |
| 3 | 70-75 | M | 1-5 | 1 | 200 mg | 3 | **Telmisartan Amlodipini**: blood pressure | 600 | 3 | 3 |
| 4 | 60-65 | M | 10-15 | 1 | 100 mg | 3 | **Sifrol ER 3 mg**: PD symptoms | 600 | 1 | 2 |
| 5 | 70-75 | M | 10-15 | 1 | 250 mg | 4 | **Sifrol ER 3 mg**: PD symptoms  **Pariet**: stomach acid | 1300 | 1 | 5 |
| 6 | 75-80 | M | 5-10 | 1 | 200 mg | 3 |  | 600 | 1 | 1 |
| 7 | 65-70 | M | 1-5 | 1 | 200 mg | 3 | **Sifrol ER 4.5 mg**: PD symptoms  **Progout**: gout  **Atacand**: blood pressure  **Flomaxtra**: unitary relief  **Lipitor**: cholesterol | 1050 | 5 | 2 |
| 8 | 50-55 | F | 1-5 | 1 | 125 mg | 3 | **Pramipexole 1.5 mg**: PD symptoms | 525 | 2 | 3 |
| 9 | 70-75 | M | 5-10 | 3 | 200 mg | 3 |  | 600 | 7 | 6 |
| 10 | 70-75 | M | 1-5 | 1 | 100 mg | 3 |  | 300 | 6 | 5 |
| 11 | 70-75 | M | 1-5 | 1 | 250 mg | 2 | **Somac**: heartburn  **Atozet**: cholesterol |  | 3 | 4 |
| 12 | 70-75 | F | 5-10 | 2 | 100 mg | 2 | **Sifrol ER 1.5 mg**: PD symptoms | 250 | 3 | 7 |
| 13 | 50-55 | M | 5-10 | 1 | 100 mg | 3 | **Azilect 1 mg**: PD symptoms | 400 | 2 | 4 |
| 14 | 55-60 | M | 5-10 | 1 | 200 mg | 4 | **Acimax**: stomach acid  **Co-diovan**: blood pressure | 800 | 2 | 2 |
| 15 | 65-70 | F | 1-5 | 1 | 50 mg | 3 | **Sifrol ER 3 mg**: PD symptoms  **Ramipril**: blood pressure | 450 | 3 | 5 |
| 16 | 70-75 | M | 5-10 | 1 | 100 mg | 3 |  | 300 | 1 | 1 |
| 17 | 60-65 | F | 5-10 | 2 | 200 mg | 3 | **Sifrol ER 1.5 mg**: PD symptoms | 750 | 1 | 5 |
| 18 | 70-75 | M | 1-5 | 1 | 250 mg | 3 | **Sifrol ER 1.5 mg**: PD symptoms | 900 | 1 | 2 |

*F, female; M, male; H&Y,* *Hoehn & Yahr;* *1, a sum of each parkinsonian medication converted into levodopa equivalent dose ^6^; 2, sum of tremor scores from the MDS-UPDRS subitems for resting tremor amplitude (item 17) and constancy (item 18) in the affected upper-limb.* **Ages and duration of disease are presented in range to limit identification of participants.*

### S2. M1 stimulation site

M1 stimulation was delivered with the coil placed tangentially to the scalp with the handle positioned backwards and rotated away from the midline by ~45º to induce a posterior-anterior current in M1 contralateral to the affected arm (*n* = 4 left arm). The optimal site of stimulation was defined as the site that elicited the largest and most consistent MEPs in the FDI ^1,7^. To find the optimal site for eliciting MEPs in FDI, numerous scalp sites were stimulated starting at C3 of the International 10-20 System and moving the coil in the anterior-posterior and lateral-medial plane in ~1 cm steps ^7-9^. The optimal site was marked as a target using the neuronavigation software and marked with water-soluble ink on a tightly fitted cap to ensure reliable coil placement throughout the experimental session.

Active motor threshold (AMT) involved an isometric contraction of 10% maximum voluntary contraction; the target EMG amplitude was presented to the participant and monitored by the experimenter in real-time using horizontal cursors on a monitor displaying ongoing EMG activity ^1,10-13^. The isometric contraction involved participants placing their forearm and hand in a pronated position on a custom-made brace and abducting their index finger to apply force against a fixed block on a custom-made brace.

#### S2.1. TMS Intensities: Data Analysis

Paired-sample *t*-tests were performed separately to examine differences between ON and OFF medication for (1) intensities used during the TMS setup procedure (AMT and SI_1mv_ intensity) and (2) the mean MEP amplitude elicited by SI_1mV_-alone trials (delivered to M1).

#### S2.2. TMS Intensities: Results

Supplementary Figure 1 shows column scatter plots of SI_1mV_ intensity (%MSO; Supplementary Figure 1A) and AMT (%MSO; Figure 1B) ON and OFF medication. There was no significant difference between sessions (ON, OFF) for SI_1mV_ intensity (*t*_17_ = 0.54, *P* = 0.599, *d* = 0.13), AMT (*t*_17_ = -0.49, *P* = 0.629, *d* = 0.12) and SI_1mV_-alone MEP amplitudes recorded from FDI (*t*_17_ = 0.30, *P* = 0.770, *d* = 0.07) or ECR (*t*_17_ = -1.24, *P* = 0.231, *d* = 0.29).

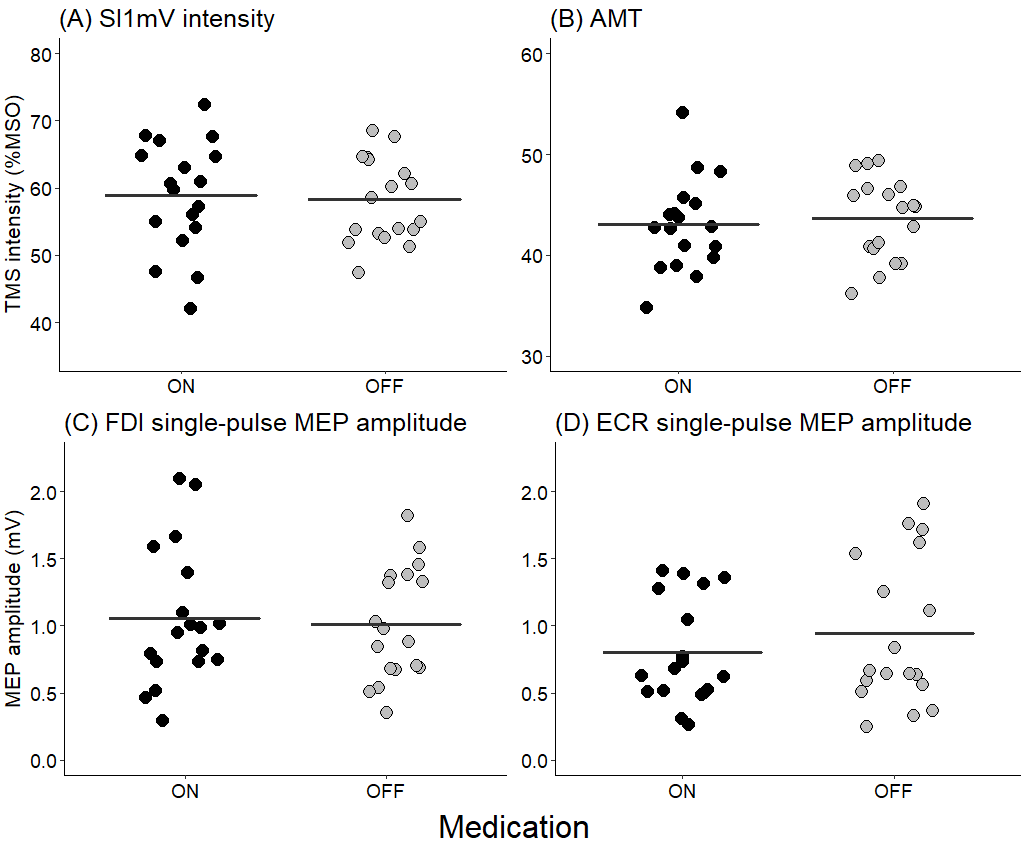

***Supplementary Figure 1.*** Column scatter graphs of (A) SI_1mV_ (%MSO) and (B) AMT (%MSO) ON (black circles) and OFF medication (grey circles). The solid lines show the mean intensity (A and B).

### S3. Custom-developed script

A custom-developed Signal script was used to trigger TMS pulses when resting EMG activity was identified between tremor bursts (see Figure 2 in the main manuscript). Prior to the TMS data collection, EMG activity from ECR was used to identify and determine a threshold for tremor bursts (in microvolts) for each individual. ECR was the target muscle because of its involvement in wrist flexion-extension tremor, and because previous research suggests that tremor bursts are more consistent in ECR than other muscles, such as FDI ^14^. During data collection, TMS pulses were triggered if ECR EMG activity was *below* an individual’s threshold (in microvolts) for a duration of 50 milliseconds (see Figure 2A in the main manuscript). If EMG activity was not below the EMG activity threshold for 50 ms, TMS was triggered after 5 seconds (±2%), indicating some EMG activity at the time of TMS delivery (see Figure 2B in the main manuscript): these trials were excluded from the analysis. ON medication, the mean number of trials excluded because of EMG activity above the defined threshold across individuals was 6 (range 0-15 across SI_1mV_-alone and dual-site trials). OFF medication, the mean number of trials excluded because of EMG activity above the defined threshold across individuals was 7 (range 0-17 across SI_1mV_-alone and dual-site trials). ON medication, individual EMG thresholds ranged from 0.011 to 0.065 mV (*M* = 0.036 mV, *SD* = 0.014 mV). OFF medication, individual thresholds ranged from 0.026 to 0.074 mV (*M* = 0.043 mV, *SD* = 0.012 mV).

### S4. Resting tremor measures

Supplementary Figure 2 shows MDS-UPDRS (Figure 2A), EMG power (Figure 2B) and acceleration power (Figure 2C) for resting tremor ON and OFF medication. Statistical analyses used to examine differences in resting tremor ON and OFF medication are reported in the main manuscript (see *Results*).

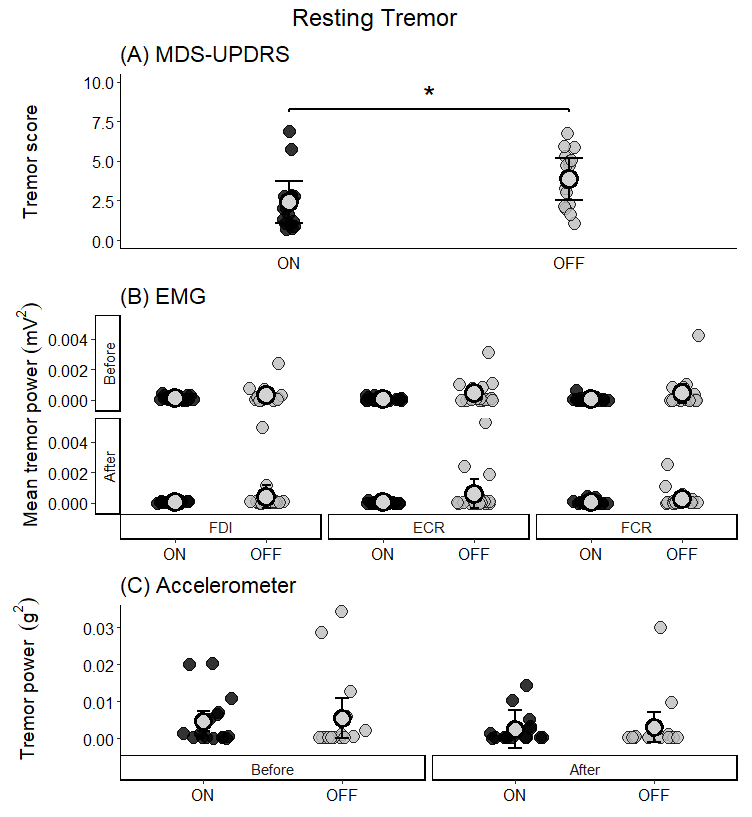

***Supplementary Figure 2.*** Resting tremor recorded from the MDS-UPDRS tremor severity score (A), EMG tremor power (mV^2^) (B), and acceleration tremor power (g^2^) ON (black circles) and OFF (grey circles) medication. EMG and acceleration were measured before and after TMS. **P < 0.05.*

### S5. Postural and action tremor

The order of the EMG and tri-axial accelerometer measures of tremor was counterbalanced across participants and sessions. The order of recording tremor type was randomised for each participant and session.

#### S5.1. Data analysis

Paired-sample *t*-tests were performed to examine differences in the MDS-UPDRS postural and action tremor severity score ON and OFF levodopa medication. GLMMs were performed separately for postural tremor and action tremor to analyse EMG tremor power (mV^2^), with fixed factors of Muscle (FDI, ECR, FCR), Medication state (ON, OFF), and Time (before TMS, after TMS), with by-subject intercept as a random effect. GLMMs were also performed separately for postural tremor and action tremor to analyse acceleration tremor power (mV^2^), with fixed factors of Medication state (ON, OFF) and Time (before TMS, after TMS), with by-subject intercept as a random effect.

#### S5.2. Results

##### S5.2.1. Postural Tremor

*MDS-UPDRS*. Supplementary Figure 3 shows postural tremor measures using the MDS-UPDRS tremor severity scores (3A), EMG tremor power (mV^2^; 3B), and acceleration tremor power (g^2^; 3C) ON and OFF medication.

A paired-sample *t*-tests showed no significant difference in postural tremor severity scores (*t*_17_ = -1.77, *P* = 0.094, *d* = 0.44) between medication states.

*EMG.* Two data points for postural tremor EMG were removed as they were clear outliers (> 12 mV^2^). The GLMM analysis for postural tremor found a significant main effect of Medication state (χ^2^(1, N = 18) = 9.35, *P* = 0.002), with higher postural tremor power during OFF compared with ON medication. There was also a significant main effect of Muscle (χ^2^(2, N = 18) = 19.66, *P* < 0.0001). Follow-up analyses showed that postural tremor power for the FDI was significantly higher than ECR (*z* = 6.13, *P* = <.0001, *d* = 1.02) and FCR (z = 4.18, *P* = 0.0001, *d* = 0.70). There was no significant difference in postural tremor power for ECR and FCR (z = -1.90, *P* = 0.173, *d* = -0.32). There were no other significant interactions nor main effects for postural tremor measured using EMG (χ^2^*s* < 1.39, *Ps* > 0.250).

*Acceleration.* Two data points for postural tremor acceleration were removed as they were clear outliers (> 0.015 g^2^). The GLMM analysis for postural tremor acceleration found no significant effect of Medication state (χ^2^(1, N = 18) = 3.50, *P* = 0.062), Time (χ^2^(1, N = 18) = 2.96, *P* = 0.085), nor interaction between Medication state and Time (χ^2^(1, N = 18) = 0.01, *P* = 0.911).

These results suggests that there was no consistent difference in postural tremor ON and OFF medication in the current sample. Given postural tremor was greater OFF than ON medication when measured with EMG, this might be the most sensitive measure for postural tremor.

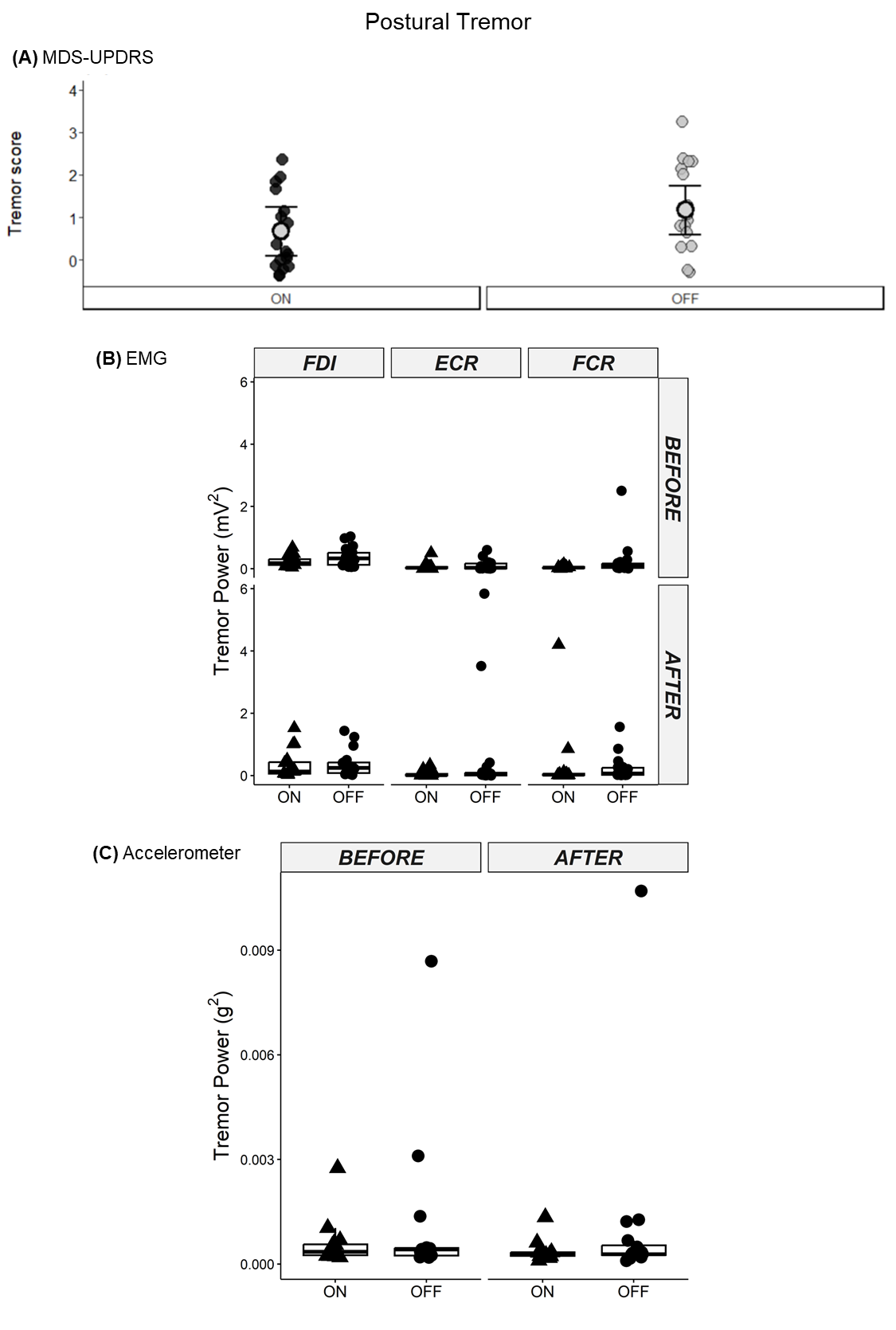

***Supplementary Figure 3.*** Postural tremor recorded from the MDS-UPDRS tremor severity score (A), EMG tremor power (mV^2^) (B), and acceleration tremor power (g^2^) ON (black circles) and OFF (grey circles) medication. EMG and acceleration were measured before and after TMS.

##### S5.2.2. Action Tremor

*MDS-UPDRS*. Supplementary Figure 4 shows action tremor measures using the MDS-UPDRS tremor severity scores (4A), EMG tremor power (mV^2^; 4B), and acceleration tremor power (g2; 4C) ON and OFF medication. A paired-sample *t*-test showed no significant difference in action tremor severity (*t*_17_ = -1.53, *P* = 0.143, *d* = 0.45) between medication states.

*EMG.* The GLMM analysis for action tremor found significant main effects of Medication state (χ^2^(1, N = 18) = 4.51, *P* = 0.033) and Muscle (χ^2^(2, N = 18) = 9.60, *P* = 0.008), which was mediated by a higher-order two-way interaction between Medication state and Muscle (χ^2^(2, N = 18) = 10.43, *P* = 0.005). Follow-up analyses showed that action tremor power for the ECR was significantly larger during OFF medication compared with ON medication (z = 3.79, *P* = 0.0001, *d* = 0.89). There were no significant differences in action tremor power between medication states for FDI (*z* = 0.47, *P* = 0.641, *d* = 0.11) and FCR (*z* = -0.58, *P* = 0.561, *d* = -0.14). There were no other significant interactions or main effects for action tremor measured using EMG (χ^2^*s* < 1.30, *Ps* > 0.254).

*Acceleration.* One data point for action tremor acceleration was removed as it was a clear outlier (> 0.3 g^2^). The GLMM analysis for postural tremor acceleration found no significant effect of Medication state (χ^2^(1, N = 18) = 1.87, *P* = 0.171), Time (χ^2^(1, N = 18) = 3.59, *P* = 0.058), nor interaction between Medication state and Time (χ^2^(1, N = 18) = 0.002, *P* = 0.963).

These results suggests that there was no consistent different in action tremor ON and OFF medication in the current sample. Given action tremor was greater OFF than ON medication when measured with EMG, this might be the most sensitive measure for action tremor.

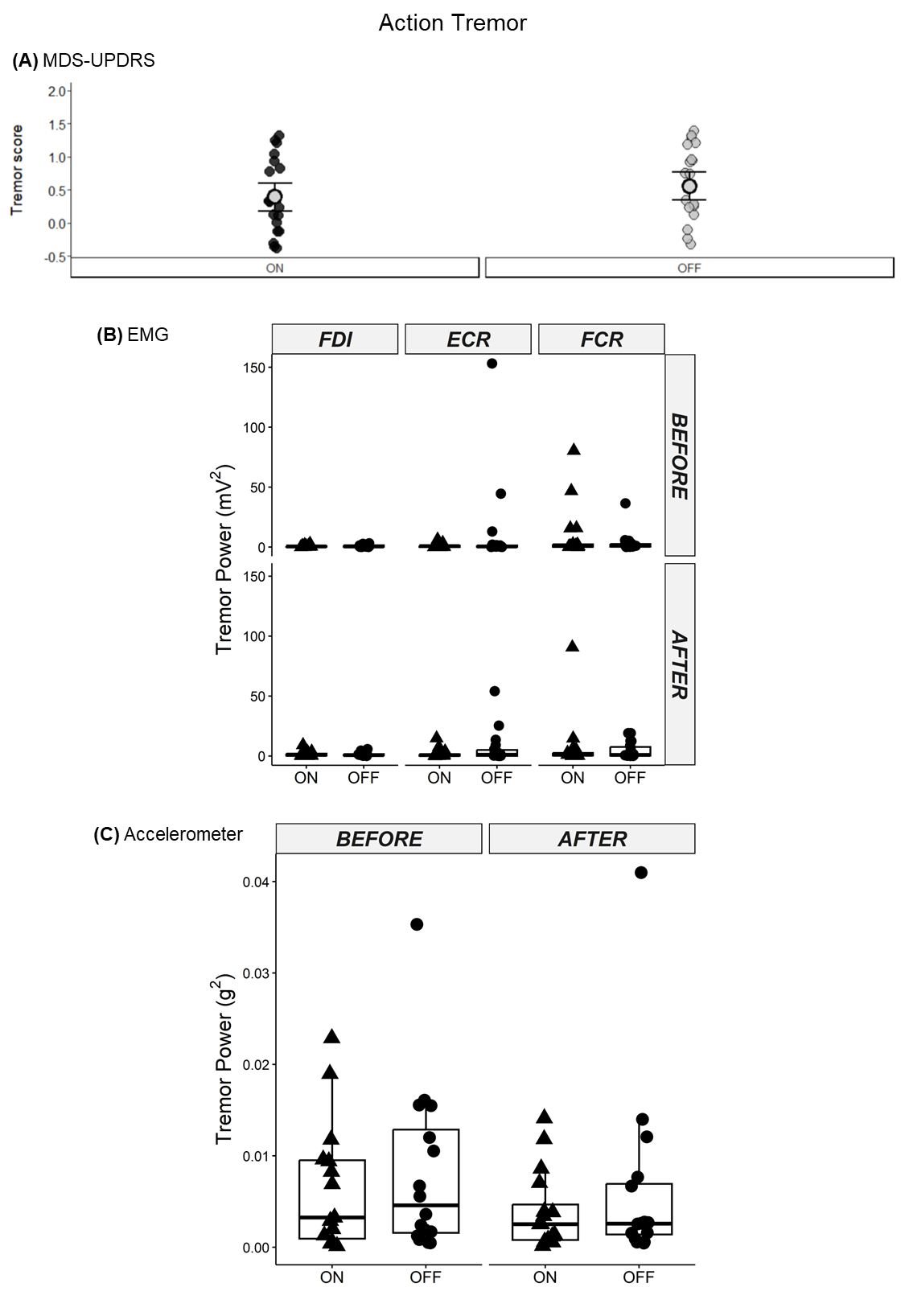

***Supplementary Figure 4.*** Action tremor recorded from the MDS-UPDRS tremor severity score (A), EMG tremor power (mV^2^) (B) and acceleration tremor power (g^2^) ON (black circles) and OFF (grey circles) medication. EMG and acceleration were measured before and after TMS.

### S6. Pre-TMS RMS as a covariate

Supplementary Figure 5 shows the effects of pre-TMS RMS as a covariate. For all trial-types, muscles, and medication states, MEP amplitudes increased as pre-TMS RMS increased. For FDI, there was no significant difference in pre-TMS RMS trends between ON and OFF medication (|*z*s| < 2.58, *P*s > 0.163), or between SI_1mV_-alone and dual-site trials (|*z*s| < 0.66, *P*s > 0.998). For ECR, the pre-TMS RMS trend for dual-site MEP amplitudes differed between medication states, with a steeper increase in dual-site MEP amplitudes with increasing pre-TMS RMS OFF than ON medication (*z* = 4.06, *P* = 0.001, *d* = 18.14). There was no significant difference in ECR pre-TMS RMS trends between ON and OFF medication for SI_1mV_-alone trials (*z* = 0.99, *P* = 0.977, *d* = 3.58), and no difference between SI_1mV_-alone and dual-site trials (|*z*s| < 2.61, *P*s > 0.152).

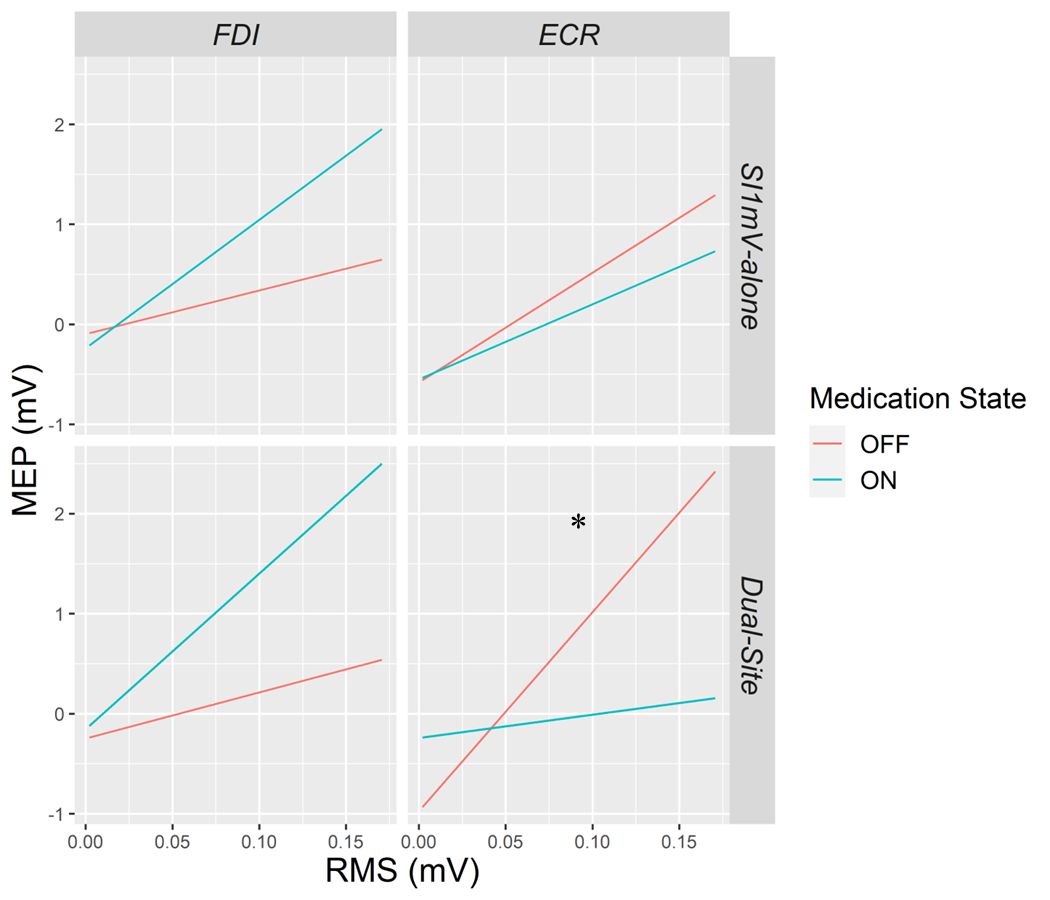

***Supplementary Figure 5.*** The estimates of the slopes of the pre-TMS RMS trend for each level of Trial-type, Muscle, and Medication state. In all conditions, as RMS increased, MEP amplitudes increased. The pre-TMS RMS trends for dual-site MEP amplitudes from the ECR were significantly different between medication states, with greater increases in MEP amplitudes from increased pre-TMS RMS (steeper pre-TMS RMS trend) OFF medication compared to ON medication. **P < 0.05.*

### S7. Relationship between resting tremor and ECR SMA-M1 connectivity

#### S7.1. Results

Supplementary Figures 6 and 7 show associations between FDI SMA-M1 connectivity ratios and resting tremor power measured using accelerometry and EMG, respectively. Positive correlations between SMA-M1 connectivity and tremor were observed in ECR ON medication (Supplementary Figure 6), with less severe tremor associated with more facilitatory ECR SMA-M1 connectivity, however, these associations were not significant after FDR adjustment.

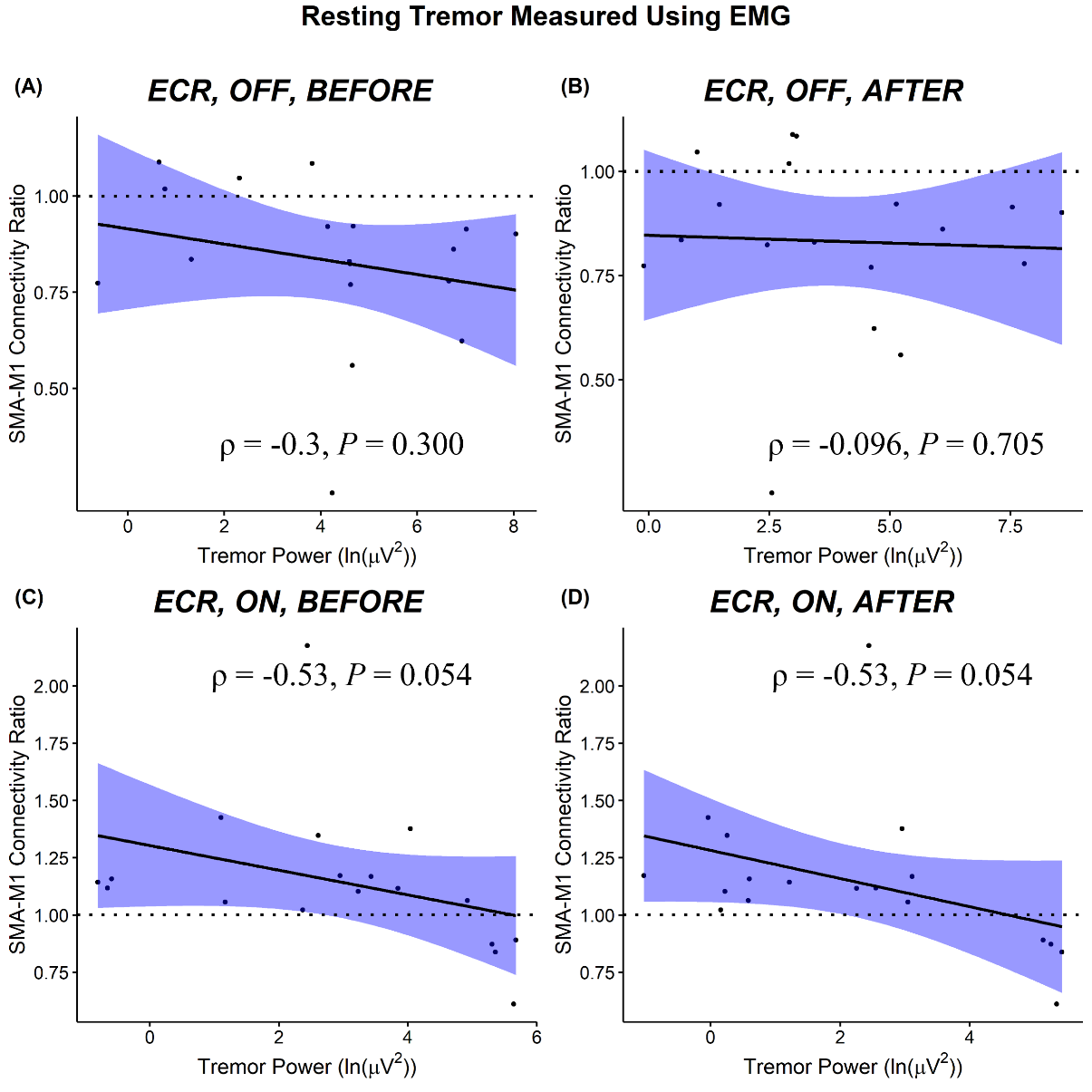

***Supplementary Figure 6.*** Scatterplots show the relationship between the magnitude of ECR SMA-M1 connectivity ratios and ECR resting tremor power measured using EMG (ln(µV^2^)). 95% confidence interval bands are shown.

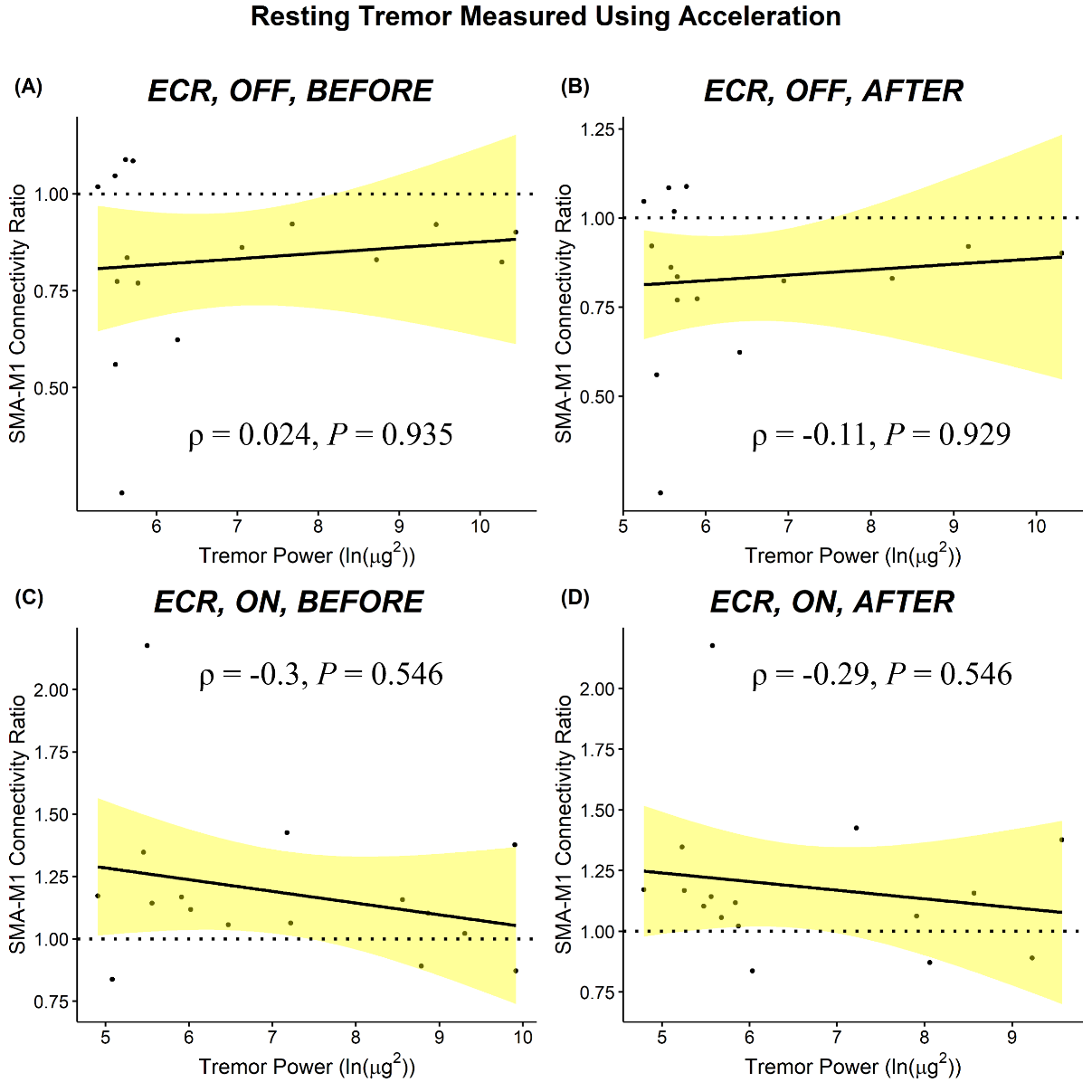

***Supplementary Figure 7.*** Scatterplots show the relationship between the magnitude of ECR SMA-M1 connectivity ratios and resting tremor power measured using acceleration (ln(µg^2^)). 95% confidence interval bands are shown.

### S8. Relationship between postural and action tremor and SMA-M1 connectivity

#### S8.1. Data analysis

As described in the main manuscript (see *SMA-M1 connectivity*), MEPs were simultaneously elicited from both FDI and ECR and, therefore, SMA-M1 connectivity was quantified separately from FDI and ECR MEPs. Exploratory Spearman’s rank correlation coefficients (ρ) were performed to examine the relationship between SMA-M1 connectivity and tremor measures (EMG, acceleration). Separate analyses were performed for medication state and tremor measures obtained before and after TMS. Correlations were performed between SMA-M1 connectivity recorded from FDI and EMG recorded from FDI and SMA-M1 connectivity recorded from ECR and EMG recorded from the ECR.

#### S8.2. Results

Supplementary Figures 8 to 15 show results for correlations between SMA-M1 connectivity (FDI, ECR) and postural and action tremor severity (EMG, acceleration). No significant correlations were found between SMA-M1 connectivity (FDI or ECR) and tremor severity, suggesting that it is unlikely that SMA-M1 connectivity is implicated in postural or action tremor.

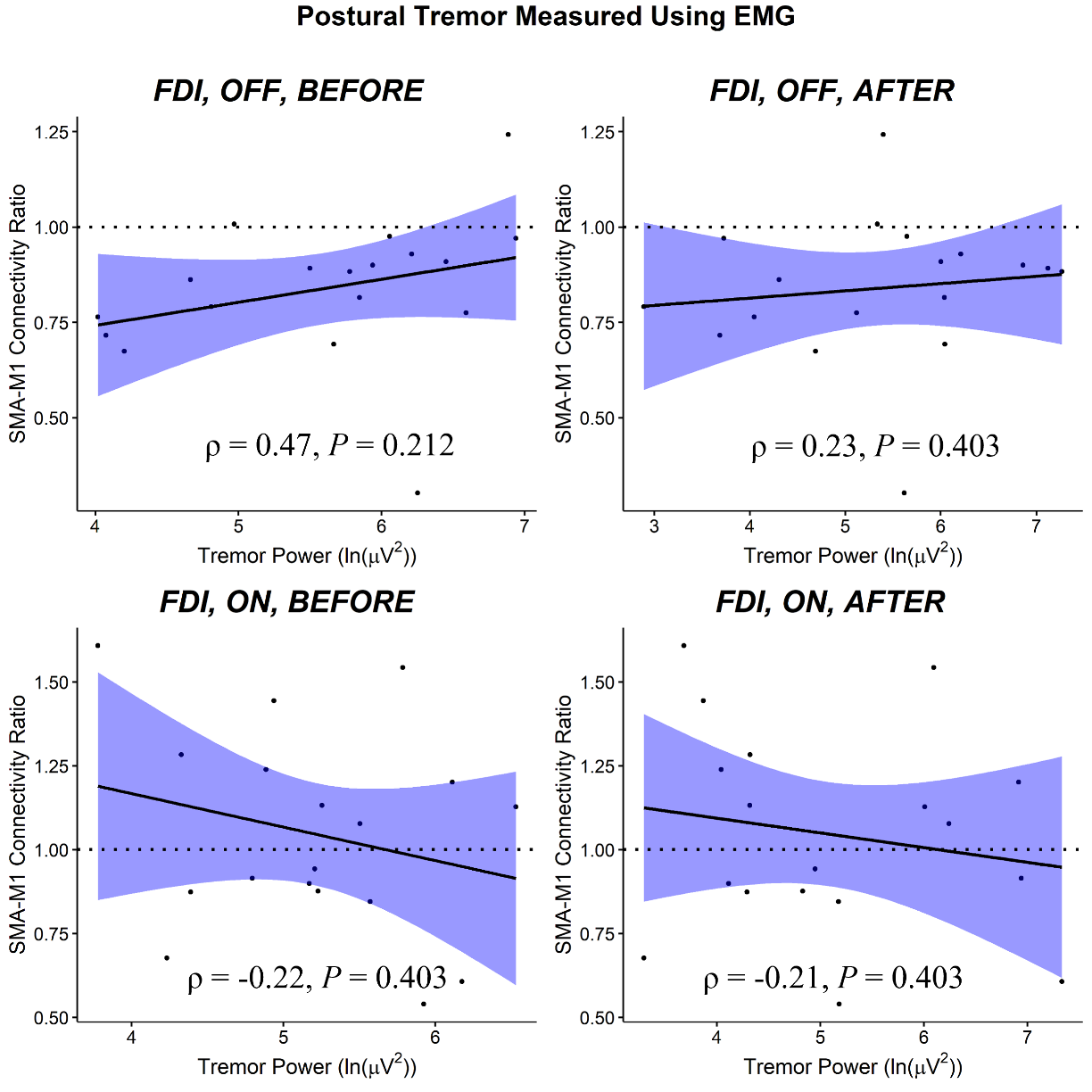

***Supplementary Figure 8.*** Scatterplots show the relationship between the magnitude of FDI SMA-M1 connectivity ratios and FDI postural tremor power measured using EMG (ln(µV^2^)). 95% confidence interval bands are shown.

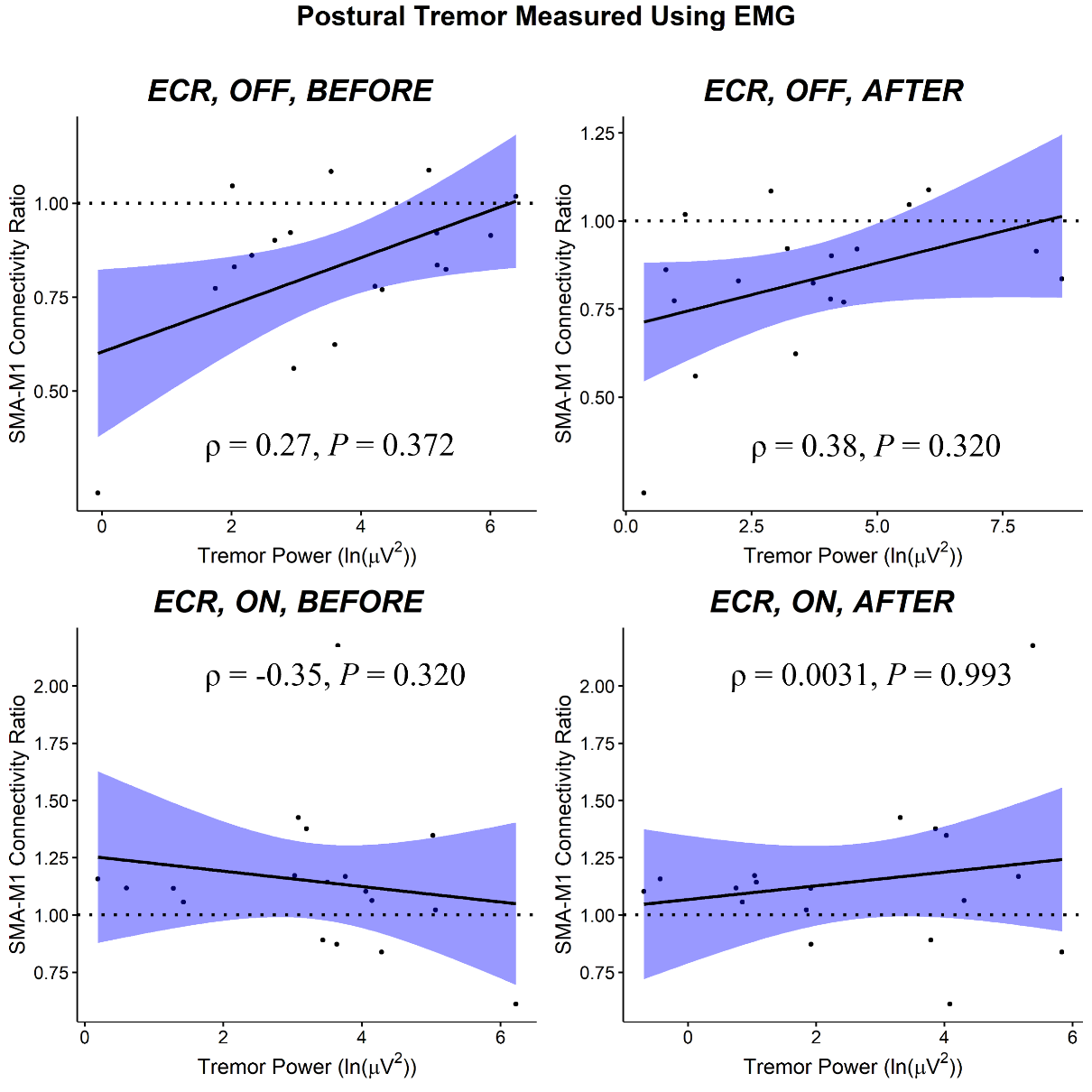

***Supplementary Figure 9.*** Scatterplots show the relationship between the magnitude of ECR SMA-M1 connectivity ratios and ECR postural tremor power measured using EMG (ln(µV^2^)). 95% confidence interval bands are shown.

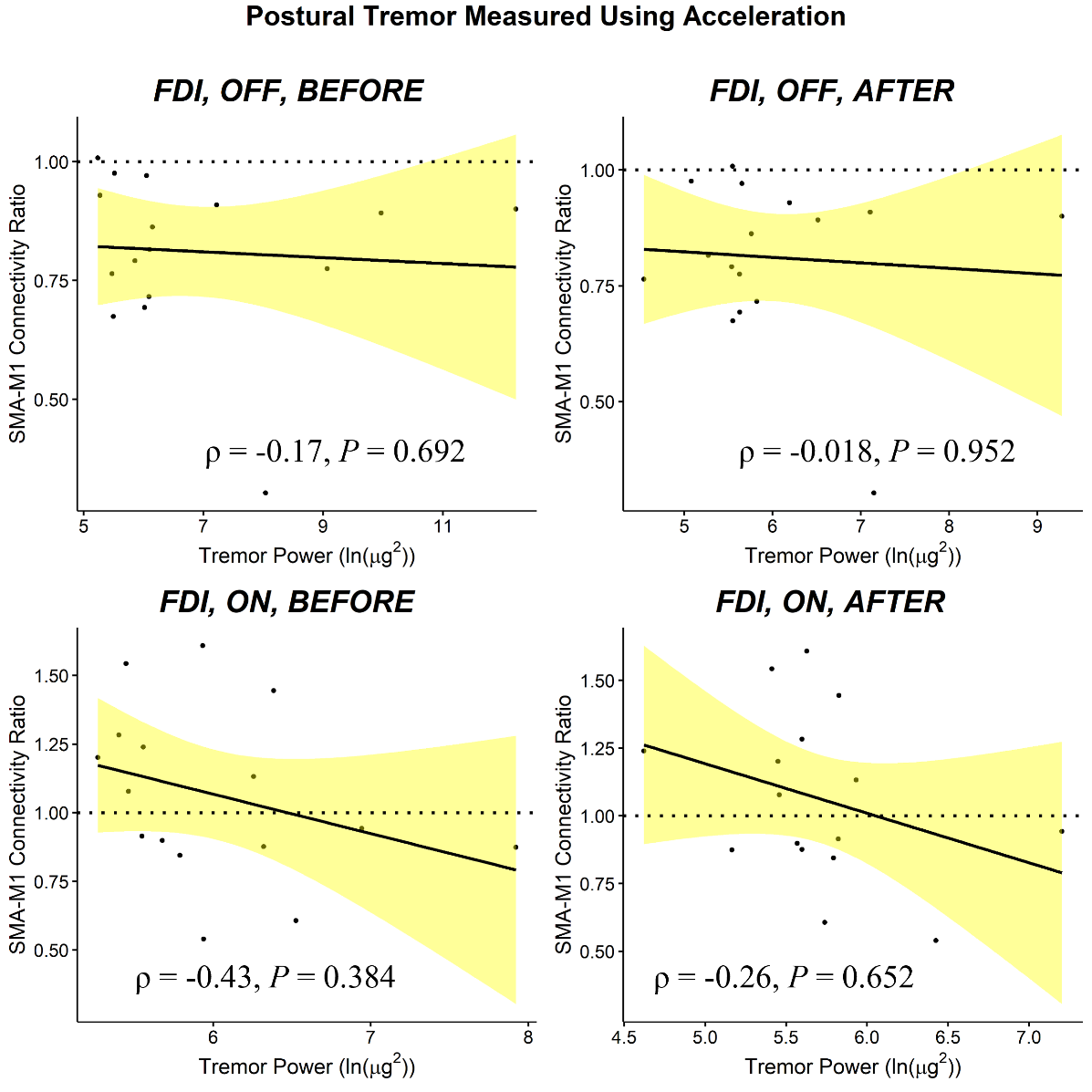

***Supplementary Figure 10.*** Scatterplots show the relationship between the magnitude of FDI SMA-M1 connectivity ratios and FDI postural tremor power measured using acceleration (ln(µg^2^)). 95% confidence interval bands are shown.

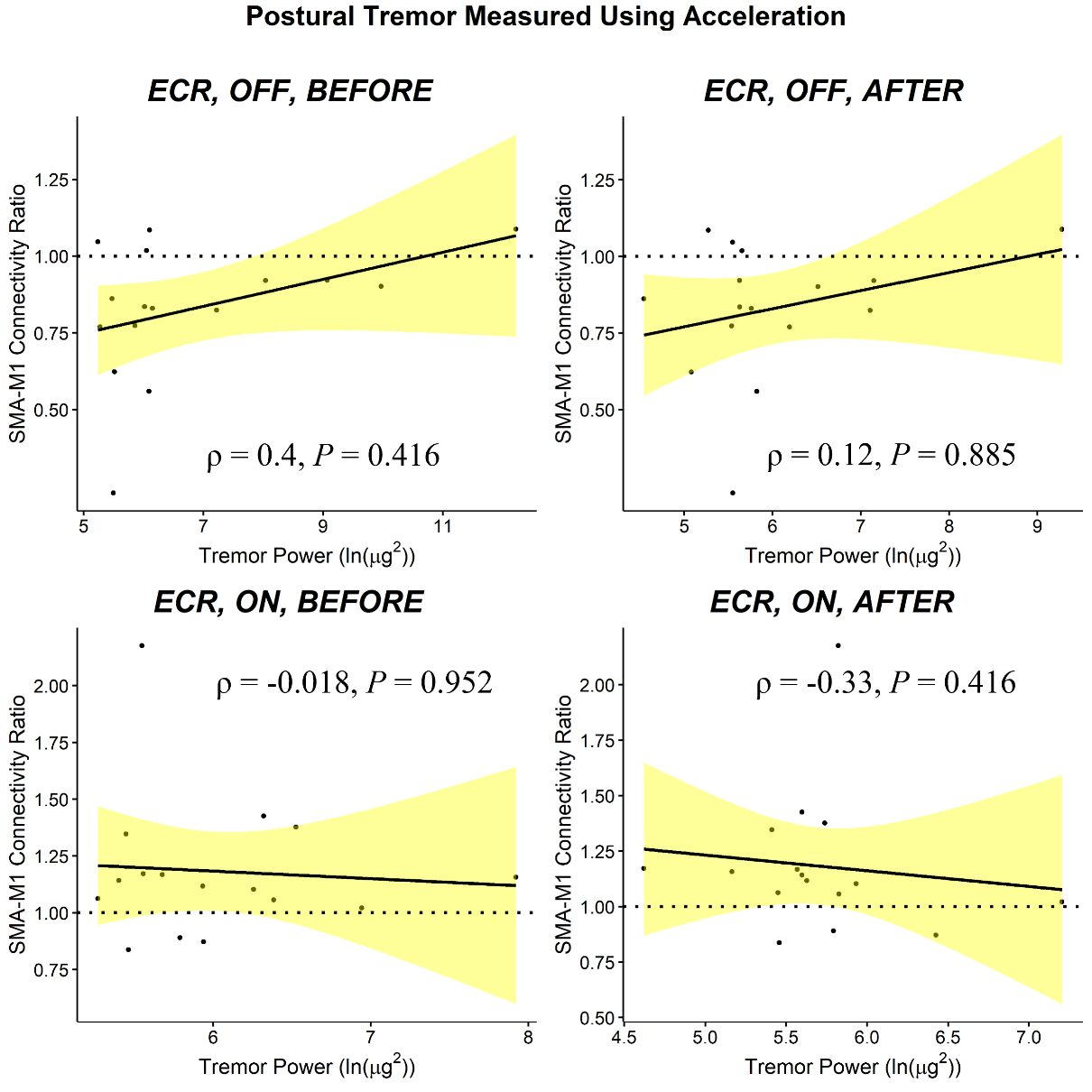

***Supplementary Figure 11.*** Scatterplots show the relationship between the magnitude of ECR SMA-M1 connectivity ratios and ECR postural tremor power measured using acceleration (ln(µg^2^)). 95% confidence interval bands are shown.

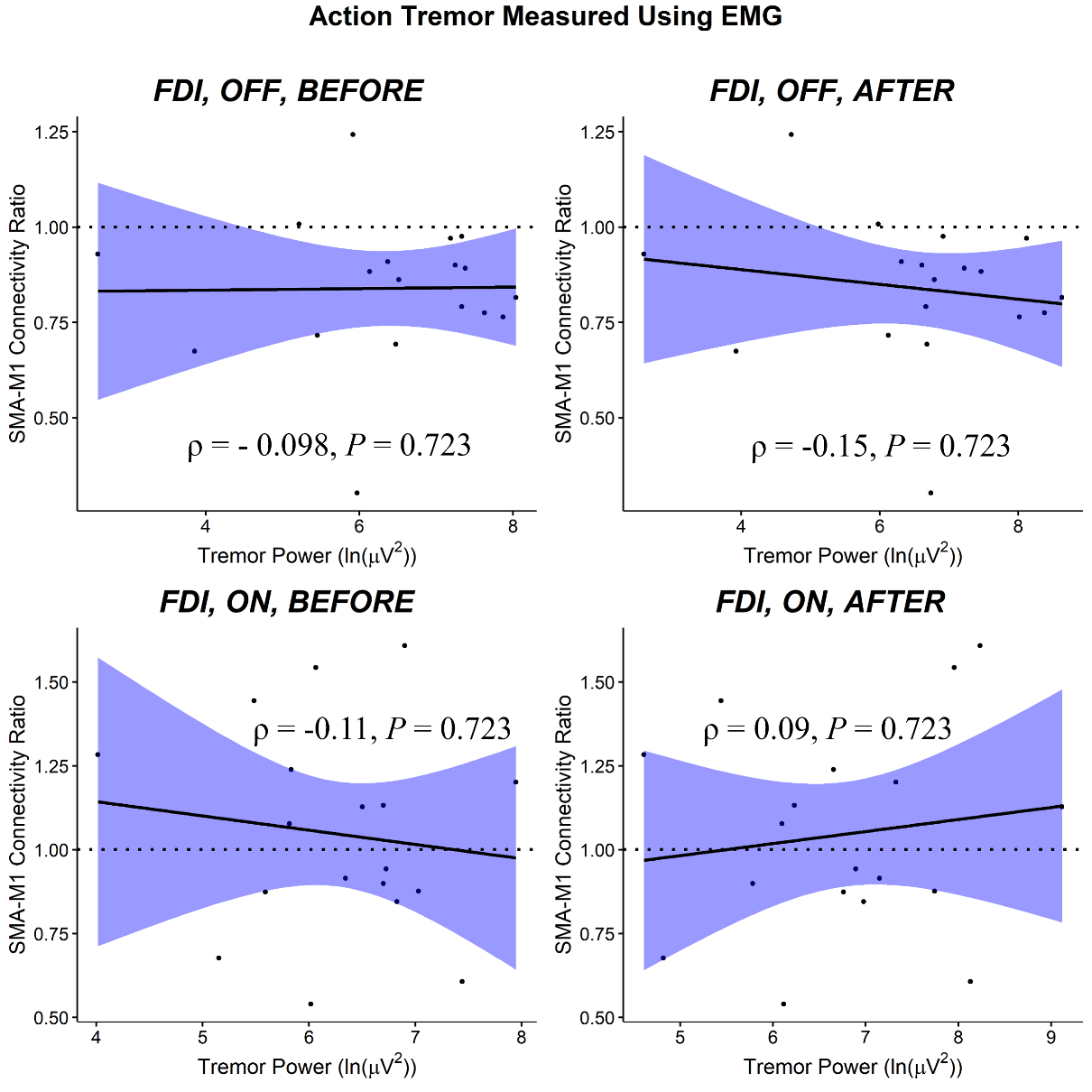

***Supplementary Figure 12.*** Scatterplots show the relationship between the magnitude of FDI SMA-M1 connectivity ratios and FDI action tremor power measured using EMG (ln(µV^2^)). 95% confidence interval bands are shown.

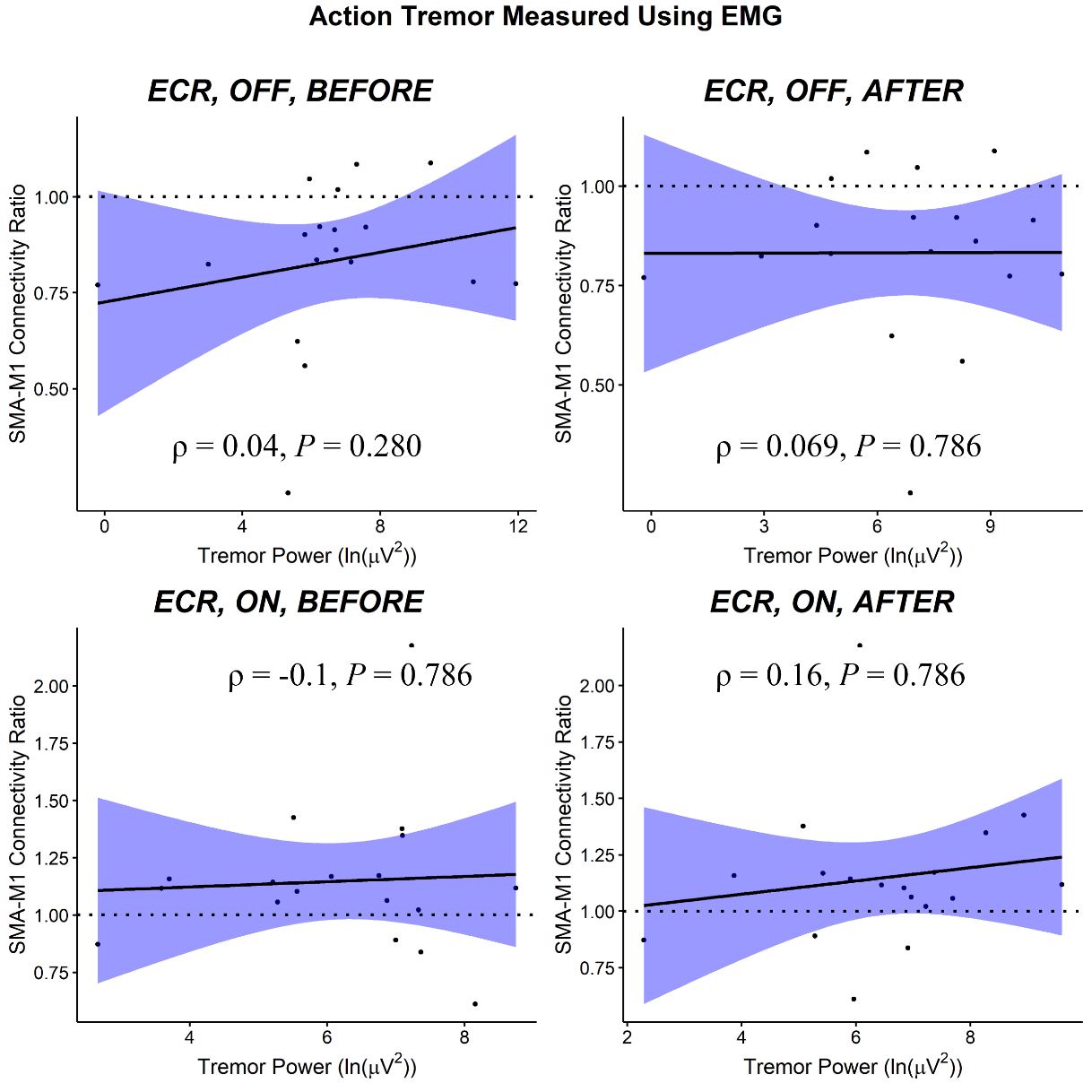

***Supplementary Figure 13.*** Scatterplots show the relationship between the magnitude of ECR SMA-M1 connectivity ratios and ECR action tremor power measured using EMG (ln(µV^2^)). 95% confidence interval bands are shown.

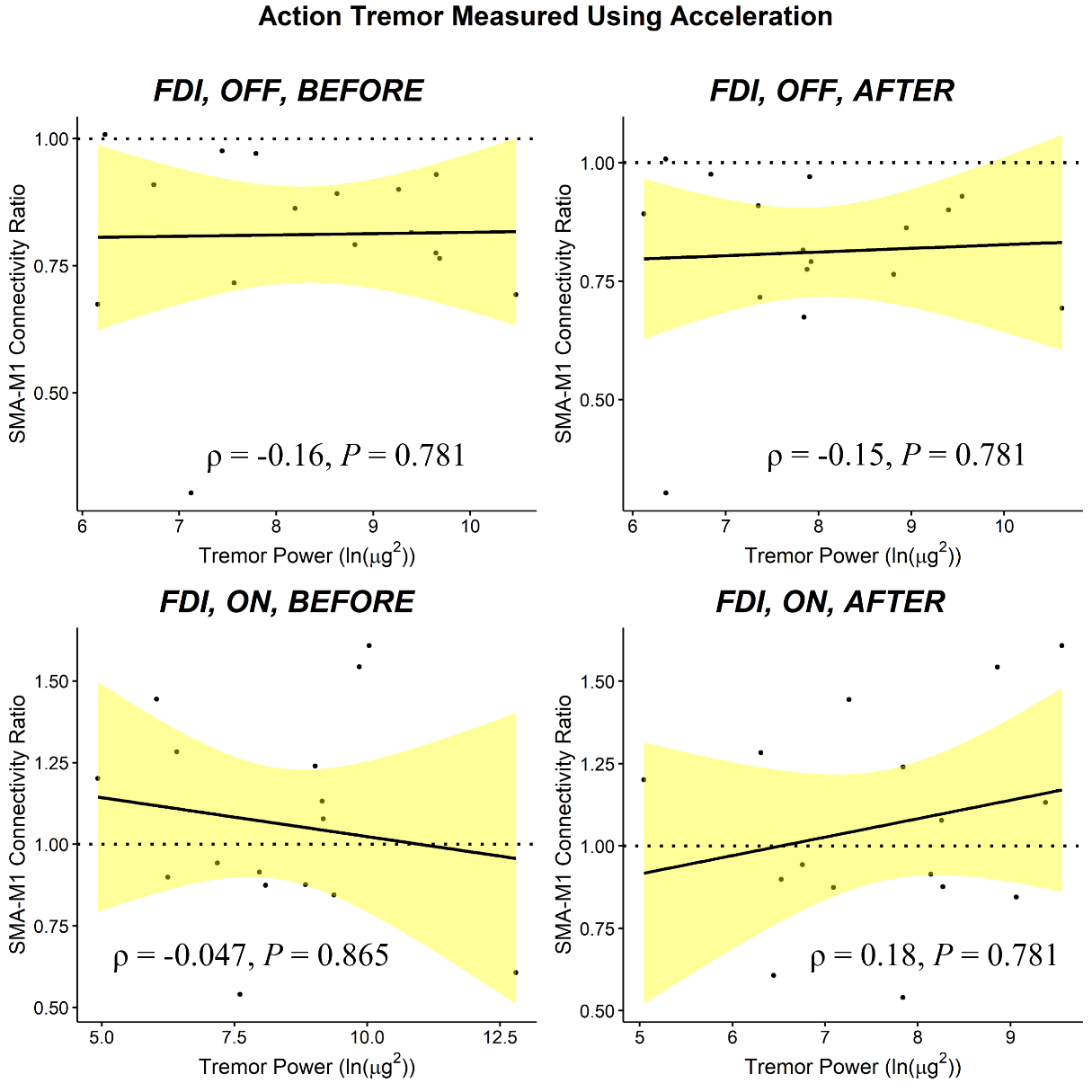

***Supplementary Figure 14.*** Scatterplots show the relationship between the magnitude of FDI SMA-M1 connectivity ratios and FDI action tremor power measured using acceleration (ln(µg^2^)). 95% confidence interval bands are shown.

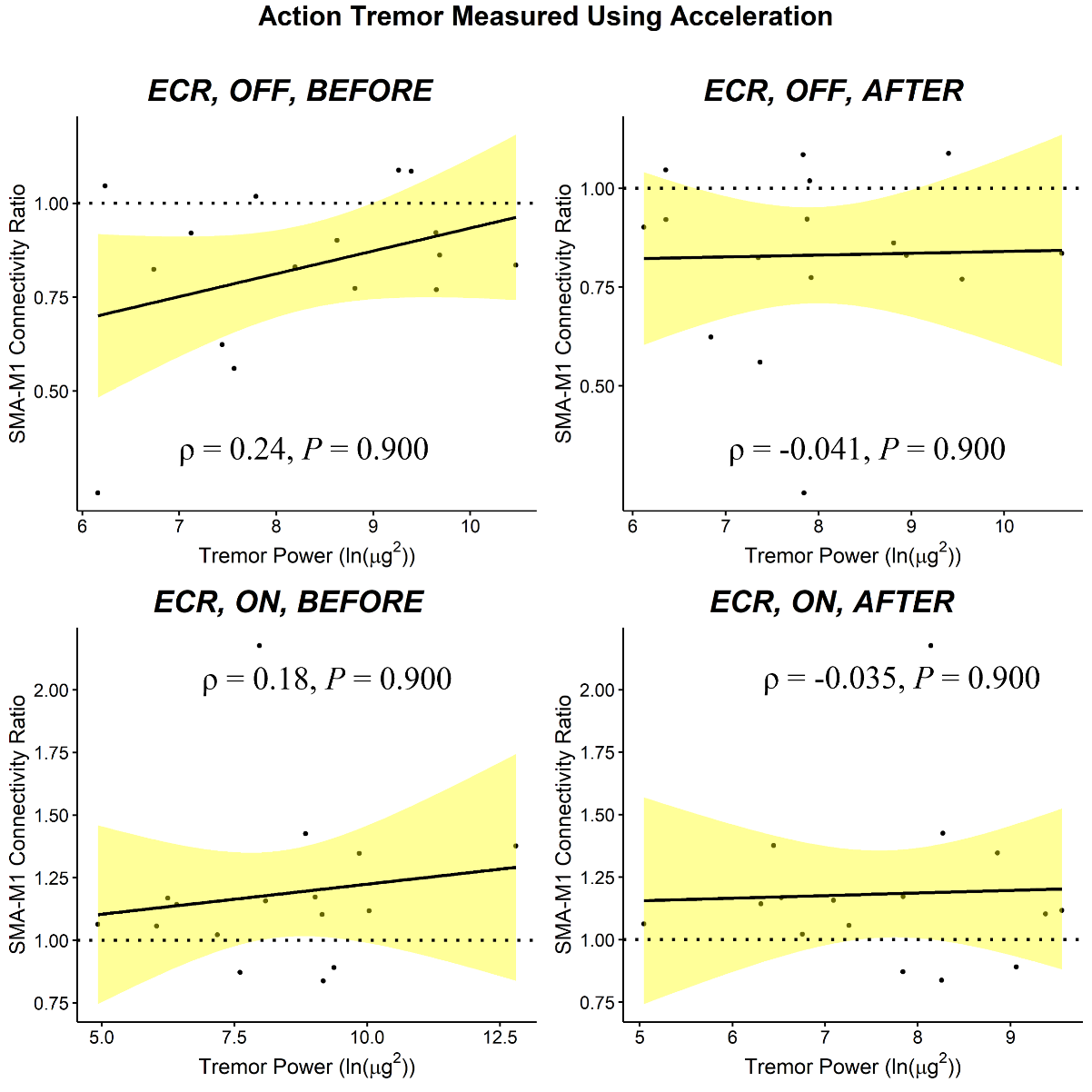

***Supplementary Figure 15.*** Scatterplots show the relationship between the magnitude of ECR SMA-M1 connectivity ratios and ECR action tremor power measured using acceleration (ln(µg^2^)). 95% confidence interval bands are shown.

### S9. Non-motor self-report questionnaires

#### S9.1. Methods

Depressive symptoms are commonly experienced in individuals with Parkinson’s disease ^16-18^. The self-report Beck Depression Inventory-II (BDI) and Patient Health Questionnaire-9 (PHQ) were used to assess depressive symptoms: a high score reflects stronger depressive symptoms ^19,20^. The self-reported questionnaires were completed in both experimental sessions and participants were asked to document depressive symptoms in the previous 14 days.

#### S9.2. Data analysis

Paired-sample *t*-tests were performed to examine differences in depression severity scores measured using the BDI and PHQ ON and OFF medication. Exploratory Pearson’s product-moment correlation coefficients were performed to examine the relationship between tremor severity (EMG, acceleration) and mean depressive scores (BDI, PHQ). Separate correlations were performed for medication state (ON, OFF), time of tremor recordings (before TMS, after TMS) and tremor type (resting, postural, action). Exploratory Pearson’s product-moment correlation coefficients were also performed to examine the relationship between depression severity scores and SMA-M1 connectivity (FDI, ECR) ON and OFF medication.

#### S9.3. Results

Paired-samples *t*-tests showed no significant difference between mean BDI depressive scores ON (*Mean* = 6.94, *SD* = 6.79) and OFF medication (*Mean* = 7.67, *SD* = 7.04) (*t*_17_ = 0.89, *P* = 0.388, *d* = 0.21) and mean PHQ depressive scores ON (*Mean* = 3.17, *SD* = 3.20) and OFF medication (*Mean* = 4.33, *SD* = 6.56) (*t*_17_ = 1.07, *P* = 0.301, *d* = 0.25). No significant associations were found between any of the tremor types and depressive mean scores on the BDI or PHQ (all *r* < 0.41, all *P* > 0.094), suggesting that it is unlikely that depressive symptoms play a role in tremor severity.

Supplementary Figures 16 and 17 show the relationship between SMA-M1 connectivity recorded from FDI (Supplementary Figure 16) and ECR (Supplementary Figure 17) and depressive scores. There were no significant associations between SMA-M1 ON medication (FDI, ECR) and depressive scores (all *r* < 0.14, all *P* > 0.578), but greater FDI SMA-M1 connectivity OFF medication was significantly associated with low depression scores on both the BDI (*r* = -0.58, *P* = 0.012) and PHQ (*r* = -0.52, *P* = 0.028); greater ECR SMA-M1 connectivity was significantly associated with low depression scores on the PHQ (*r* = -0.51, *P* = 0.031), but just failed to reach statistical significance for depressive scores on the BDI (*r* = -0.47, *P* = 0.051).

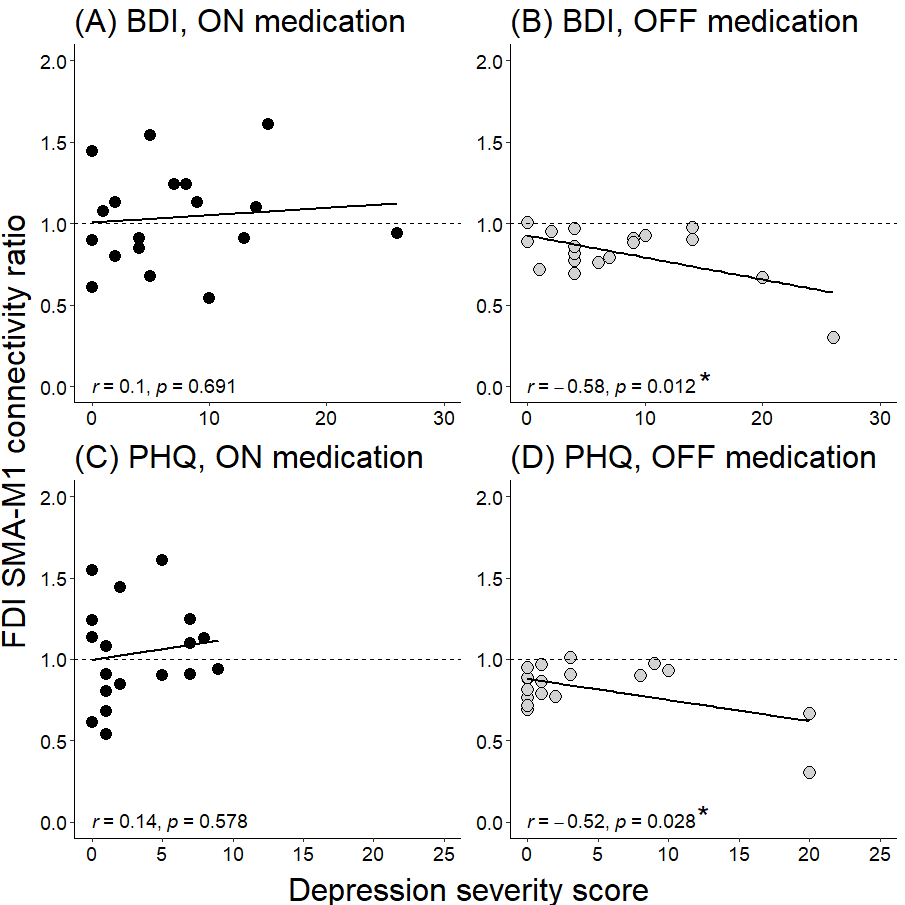

***Supplementary Figure 16.*** Scatterplots show the relationship between the magnitude of FDI SMA-M1 connectivity ratios and depressive scores recorded from the BDI (A-B) and PHQ (C-D) ON (black circles) and OFF medication (grey circles).

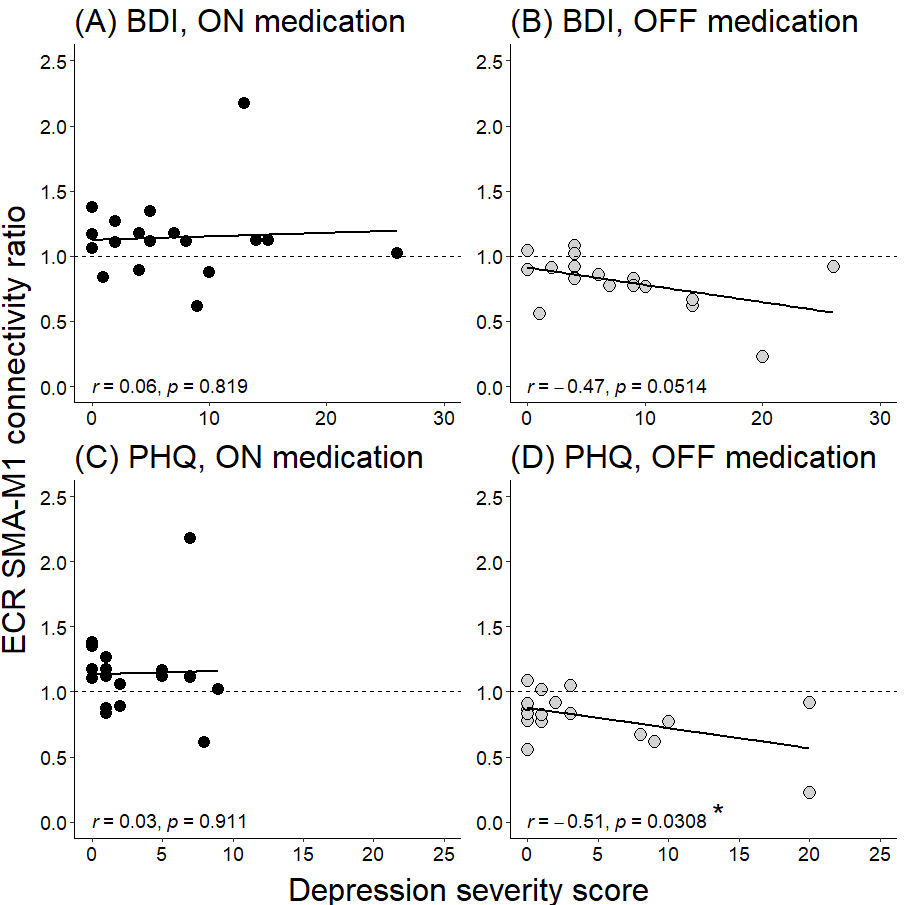

***Supplementary Figure 17.*** Scatterplots show the relationship between the magnitude of ECR SMA-M1 connectivity ratios and depressive scores recorded from the Beck Depression Index (BDI; A-B) and Patient Health Questionnaire (PHQ; C-D) ON (black circles) and OFF medication (grey circles).

#### S9.4. Discussion

Self-reported psychological depression scores from the BDI and PHQ were not significantly different ON and OFF medication, suggesting that the minimal depressive symptoms experienced in the current sample was similar both ON and OFF medication. There were no significant associations between depression scores (BDI, PHQ) and tremor recorded using EMG (FDI, ECR) or the accelerometer, suggesting that depressive symptoms were not associated with tremor. Previous research suggests that higher levels of depression was associated with greater severity in motor symptoms measured as the sum of UPDRS scores ^21^, but less is known about the influence of depression on resting tremor severity. In the current study, mean depressive scores recorded from both self-report questionnaires indicated that at the group level, there was minimal depressive symptoms. However, two individuals reported moderate or severe depressive scores, and these individuals presented with some of the strongest tremor recorded using EMG and acceleration. It is possible that tremor severity would be associated with more sever depressive symptoms or clinically diagnosed depression. However, this is speculative, and future research should examine a broader range of resting tremor severity and depression levels to comprehensively examine the relationship between the two factors.

This is the first report showing that strong inhibition of SMA on M1 was associated with high depressive scores in people with PD. It is not clear why SMA-M1 inhibition OFF medication was associated with depression, but it is tempting to speculate that this might be due to the progressive decline of serotonin neurons in the raphe nuclei ^22-26^. These neurons have strong connections with the frontal cortex and the basal-ganglia-thalamo-cortical circuit via the striatum ^27,28^. Although speculative, a decline of serotonin neurons might contribute to SMA-M1 inhibition and increase depressive symptoms. It is worth noting that most individuals in the current study presented with minimal depressive symptoms OFF medication, but two individuals presented with either moderate or severe depression, and these individuals showed the strongest SMA-M1 inhibition. Indeed, these two individuals were driving significant associations between SMA-M1 connectivity and depression and, therefore, these results should be interpreted with caution. Future research should examine SMA-M1 connectivity using dual-site TMS in people with tremor-dominant PD with and without clinically diagnosed depression to better understand the role of depression in cortico-cortical connectivity.

Depression scores were not associated with SMA-M1 connectivity ON medication. PD depression has been shown to be modulated by dopaminergic medication, with stronger depressive symptoms occurring OFF than ON medication ^29^. Short-term changes in anxiety and depressive symptoms can occur, termed non-motor fluctuations, with emergence during OFF periods and improvement during ON periods ^29^. In the current study, it is possible that dopamine medication restored serotonin mechanisms underpinning SMA-M1 connectivity and depression, leading to no associations between SMA-M1 connectivity and depression. However, as this is speculative, future research should use positron emission tomography to examine the influence of reduced serotonin on the basal ganglia-thalamo-SMA-M1 network ON and OFF medication, and any association with depression.
